# Delays in accessing high-quality care for newborns in East Africa: An analysis of survey data in Malawi, Mozambique, and Tanzania

**DOI:** 10.1101/2023.08.28.23294736

**Authors:** Lori Niehaus, Ashley Sheffel, Henry Kalter, Agbessi Amouzou, Alain Koffi, Melinda K. Munos

## Abstract

**Background:** Despite the existence of evidence-based interventions, substantial progress in reducing neonatal mortality is lagging, indicating that small and sick newborns (SSNs) are likely not receiving the care they require to survive and thrive. The “three delays model” provides a framework for understanding the challenges in accessing care for SSNs. However, the extent to which each of the delays impacts access to care for SSNs is not well-understood. To fill this evidence gap, we explored the impact of each of the three delays on access to care for SSNs in Malawi, Mozambique, and Tanzania.

**Methods:** Secondary analyses of data from three different surveys served as the foundation of this study. To understand the impact of delays in the decision to seek care (delay 1) and the ability to reach an appropriate point of care (delay 2), we investigated time trends in place of birth disaggregated by facility type and explored care-seeking behaviors for newborns who died. To understand the impact of delays accessing high-quality care after reaching a facility (delay 3), we measured facility readiness to manage care for SSNs and used this measure to adjust institutional delivery coverage for SSN care readiness.

**Findings:** Coverage of institutional deliveries was substantially lower after adjusting for facility readiness to manage SSN care, with decreases of 30 percentage points (pp) in Malawi, 14pp in Mozambique, and 24pp in Tanzania. While trends suggest more SSNs are born in facilities, substantial gaps remain in facilities’ capacities to provide lifesaving interventions. In addition, exploration of care-seeking pathways revealed that a substantial proportion of newborn deaths occurred outside of health facilities, indicating barriers in the decision to seek care or ability to reach an appropriate source of care may also prevent SSNs from receiving these interventions.

**Conclusions:** Investments are needed to overcome delays in accessing high quality care for the most vulnerable newborns – those who are born small or sick. As more mothers and newborns access health services in low- and middle-income countries, ensuring that life-saving interventions for SSNs are available at the locations where newborns are born and seek care after birth is critical.

## Background

While globally, there has been great success in increasing child survival, the decline in under-five mortality over the last three decades has not been uniform among ages. In 2020, nearly half (47%) of all under-five deaths occurred during the neonatal period (first 27 days of life), up from just 40% in 1990 (1). In addition, deaths within the neonatal period are not evenly distributed by age, with nearly three-fourths of all newborn deaths occurring within the first week of life (1). Despite widespread recognition that neonatal survival must be prioritized, 61 countries are not on track to meet their neonatal mortality rate (NMR) target for Sustainable Development Goal (SDG) three – no more than 12 neonatal deaths per 1000 live births by 2030 (2).

Notwithstanding these setbacks, effective interventions exist to save newborn lives. The Every Newborn Action Plan (ENAP) – endorsed by 194 United Nations (UN) member states and over 80 partners – identified two evidence-based packages of care with the highest impact for ending preventable newborn deaths: labor, delivery and first week of life care; and small and sick newborn care (3). Small and sick newborns (SSNs) – those weighing less than 2,500 grams at birth (including preterm and small for gestational age newborns) or having any medical or surgical condition (4) – are at the highest risk of mortality (5–7). However, a number of effective interventions for the care of SSNs have been associated with significant reductions in neonatal mortality risk (8–12). These life-saving interventions largely require facility-based care (8,9), and thus it is critical that SSNs are able to access timely, high-quality care at heath facilities.

Despite the existence of evidence-based interventions, substantial progress in reducing neonatal mortality has not been made, indicating that SSNs are likely not receiving the care they require to survive and thrive when and where they need it. The Three Delays Model provides a useful framework for understanding the challenges in accessing care for SSNs. The model was first proposed by Thaddeus & Maine in 1994 to understand the proximal causes of maternal mortality and access to care (13). It breaks down barriers in the care-seeking process into three categories: 1) delays in the decision to seek care, 2) delays in reaching an appropriate source of care (usually, a health facility), and 3) delays in receipt of adequate and appropriate care after reaching the facility. Understanding the relative impact of these delays on access to health services for newborns can help guide context-specific policies and interventions.

The extent to which each of these delays impacts access to care for SSNs is not well-understood, and there is minimal information available on care-seeking behaviors for newborns. Previous studies have explored increases in institutional delivery coverage (14–17) that suggest more newborns are reaching health facilities during the critical window of labor, delivery and the first week of life, thus overcoming delays 1 and 2 immediately after birth. However, few studies include detailed information on the types of facilities, including level (e.g., hospitals or primary facilities) and managing authority (e.g., government or private), from which care is sought. A more granular understanding of these facility types, and more information on care-seeking behaviors for newborns after birth, would identify delays in reaching the appropriate level of care and allow for more targeted intervention strategies.

Additionally, understanding only where newborns seek care is insufficient to assess whether they were able to access necessary, high-quality services after reaching the appropriate facility (i.e., whether they experienced delay 3). Many factors may influence receipt of adequate care for SSNs in health facilities, including inadequate staffing, poor availability of equipment, and low health worker competency to deliver SSN care interventions (18). Quality-adjusted coverage measures, also known as effective coverage, can be used to provide evidence to assess whether newborns who reached health facilities may still face delays in receipt of high-quality services, due to poor newborn health service availability or service provision (19,20). Poor facility readiness, or the availability of inputs at a health facility that are required to deliver newborn health services, may result in delays to SSNs receiving the care they need, but we are unaware of any studies that have explicitly adjusted coverage for facility readiness to provide lifesaving interventions for SSNs.

In an effort to fill these evidence gaps, we analyzed available survey and social autopsy data from Malawi, Mozambique, and Tanzania to better understand the impact of each of the three delays on access to high quality care for SSNs. To understand the impact of delays in the decision to seek care (delay 1) and the ability to reach an appropriate point of care (delay 2), we investigated time trends in place of birth disaggregated by facility type (level and managing authority) and explored care-seeking behaviors for newborns who died. To understand the potential of newborn service input availability to lead to delays in SSN care after reaching a facility (delay 3), we measured overall and intervention-specific readiness to manage care for SSNs disaggregated by facility type and used the resulting scores to adjust institutional delivery coverage for SSN care readiness. Together, these analyses aimed to explore where interventions targeting neonatal survival may be most impactful.

## Methods

### Country selection

Three countries in East Africa were selected for this analysis based on their neonatal mortality burdens and verbal and social autopsy data availability: Malawi, Mozambique, and Tanzania. All three have neonatal mortality rates above the global average of 17 deaths per 1,000 live births – 19 per 1,000 in Malawi, 28 per 1000 in Mozambique, and 20 per 1,000 in Tanzania (2021) – and thus far have not demonstrated sufficient progress to meet the SDG NMR target for 2030 (Target 3.2) (1).

### Data sources

Secondary analyses of data from three types of surveys served as the foundation of this study: 1) Household surveys (HH), 2) Health facility assessments (HFA), and 3) Verbal and social autopsy interviews (VASA). Information on each data source and its use in the analysis is available in **Table 1**.

**Table 1.** Data sources.

| Survey Type | Country | Source | Use in analysis |
| --- | --- | --- | --- |
| **Household survey (HH)** | Malawi | DHS 2000, DHS 2004, MICS 2006, DHS 2010, MICS 2013-14, DHS 2015-16 | - Time trends of place of birth (all)<br>- Service-contact and readiness-adjusted institutional delivery coverage (Malawi DHS 2015-16, Mozambique AIS 2015, Tanzania DHS 2015-16) |
|  | Mozambique | DHS 2003, DHS 2011, AIS 2015 |  |
|  | Tanzania | DHS 2004-05, DHS 2010, AIS/MIS 2011-12, DHS 2015-16 |  |
| **Health facility assessment (HFA)** | Malawi | SPA 2013-14 | - Facility readiness scores<br>- Service contact and readiness-adjusted institutional delivery coverage |
|  | Mozambique | SARA 2018 |  |
|  | Tanzania | SPA 2014-15 |  |
| **Verbal and social autopsy survey (VASA)** | Malawi | CHERG<sup>a</sup> Stillbirth, Neonatal, and Child VASA questionnaire, 2013 | - Care-seeking behaviors for newborns who died |
|  | Mozambique | COMSA<sup>b</sup> Stillbirth, Neonatal, Child and Adult VASA questionnaire, 2018-2020 |  |
|  | Tanzania | CHERG Stillbirth, Neonatal, and Child VASA questionnaire, 2017/18 |  |

<sup>a</sup> The Child Health Epidemiology and Reference Group (CHERG) was a technical advisory group that worked with international health institutions to improve “evidence on the causes and determinants of maternal, neonatal, and child morbidity and mortality, on intervention coverage, and on the effectiveness of interventions to inform and influence global priorities and programs”(42).
<sup>b</sup> Countrywide Mortality Surveillance for Action (COMSA) is a national sample registration system for mortality and causes of death monitoring implemented by through a collaboration between the Mozambique National Institute of Health and the Institute for International Programs at Johns Hopkins Bloomberg School of Public Health. Data collection has been ongoing since roll-out began in March 2018 (41).

HH data and questionnaires, specifically Demographic and Heath Surveys (DHS), AIDS Indicator Surveys (AIS), and Multiple Indicator Cluster Surveys (MICS), are publicly available through the DHS Program (21) (DHS and AIS) or from United Nations’ Children’s Fund (UNICEF) (22) (MICS). HFA data access varied by country, with Service Provision Assessments (SPA) used for Malawi and Tanzania publicly available through the DHS Program (21), and the Mozambique 2018 Service Availability and Readiness Assessment (SARA) accessed with permission from the Mozambique World Health Organization (WHO) Country Office and the Mozambique National Institute of Health (INS). VASA de-identified data and questionnaires were accessed with permission from the primary research team at Johns Hopkins University.

All included household surveys were nationally representative. The Malawi 2013-14 SPA (23) and the Mozambique 2018 SARA (24) were censuses of all formal-sector health facilities in each country. The Tanzania 2014-15 SPA (25) was a sample of facilities stratified by region and facility type (level and managing authority). VASA data for Mozambique and Tanzania were nationally representative, whereas data for Malawi was collected in two districts – Balaka and Salima – located in the South and Central regions of Malawi and was representative of those two districts. Additional information on representativeness and sample sizes of surveys can be found in the supplementary material (**Table A-1**), and additional information on sampling design and data collection procedures is contained within the survey reports for HHs and HFAs (23–38) and in publications by the primary research teams for VASA (39–41).

### Defining the Three Delays to Newborn Care

We adapted the original framework on the Three Delays to define the delays in access to care for newborns after birth, following the same approach that has been used in previous studies focused on this population (Table 2) (43–45).

**Table 2.**
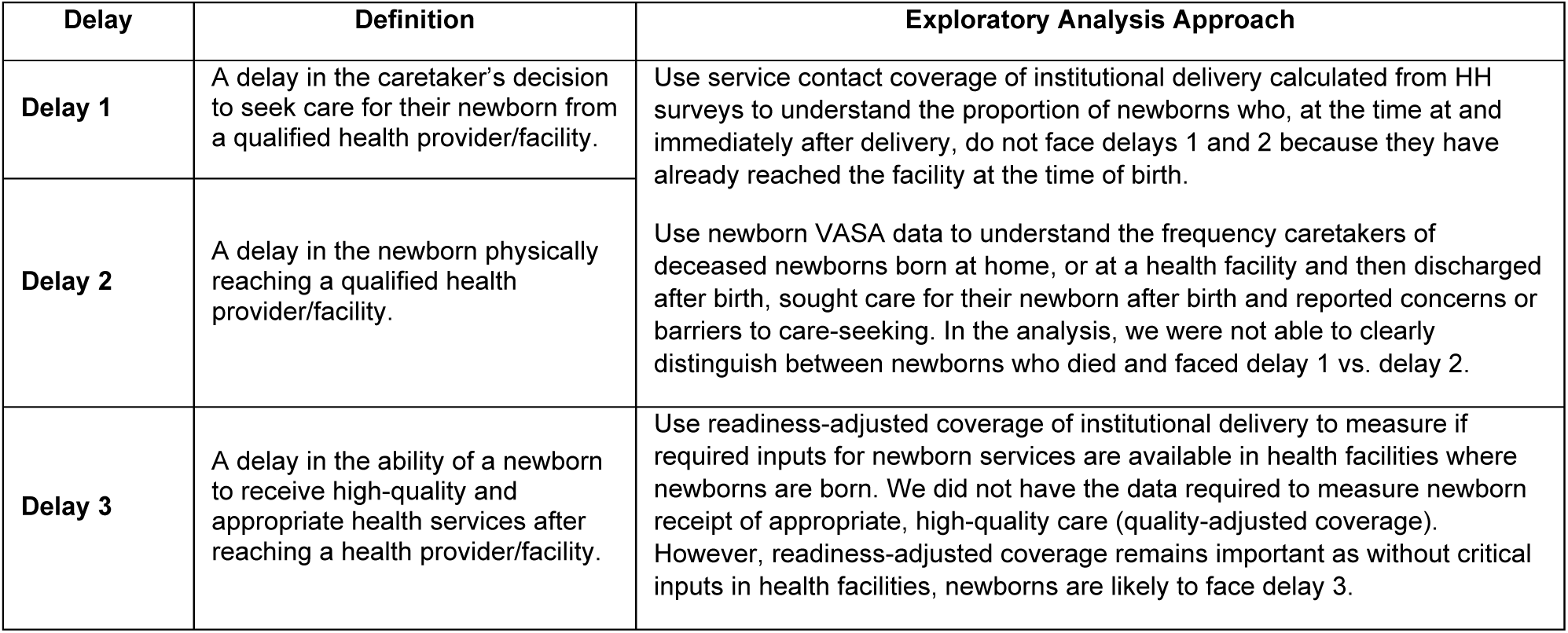
Definitions of delays in access to newborn care.

### Data analysis

#### National time trends in place of birth

In each country, we mapped place of birth response options in HH questionnaires over time to generate a set of health facility categories for which it was possible to generate time trends for place of birth. Across analyses, we defined ‘facility type’ by the facility level (e.g., secondary/hospital, primary/health center, health post) and managing authority (e.g., government, private, religious). However, a few household surveys collected less detailed information on facility level and managing authority, leading to broader facility type categories in those settings.

We first calculated from each survey the service contact coverage of institutional delivery defined as the proportion of the most recent live births of interviewed women in the 2, 3, or 5 years preceding the survey that took place in a health facility of any type ^1^. Next, for each survey year, we disaggregated institutional deliveries by level alone (primary vs. secondary) and by more detailed health facility type categories as described above. All analyses accounted for the HH survey design (clustering, stratification, and survey weights).

#### Care-seeking behaviors for newborns who died

We used VASA data to understand the care-seeking itinerary for newborns who died (**Figure 1**). We first disaggregated newborns by their place of birth (hospital, primary health facility, or community (including en route births)). Then, we calculated the proportion of newborns who were born in a health facility and died before discharge from the facility in which they were born. Among newborns born in facilities who were discharged before death and newborns born in the community, we determined the proportion who sought care from a health facility, respectively, after discharge or birth, and of these newborns, the proportions who ultimately died at a hospital, primary health facility, or in the community. All newborns of caretakers who sought care after a home birth or discharge from the health facility where they gave birth were treated equally, regardless of the number of facilities from which they sought care. We also calculated the proportion of newborns born in the community who never interacted with the health system (i.e., never sought facility-based care for any reason).

**Figure 1.**
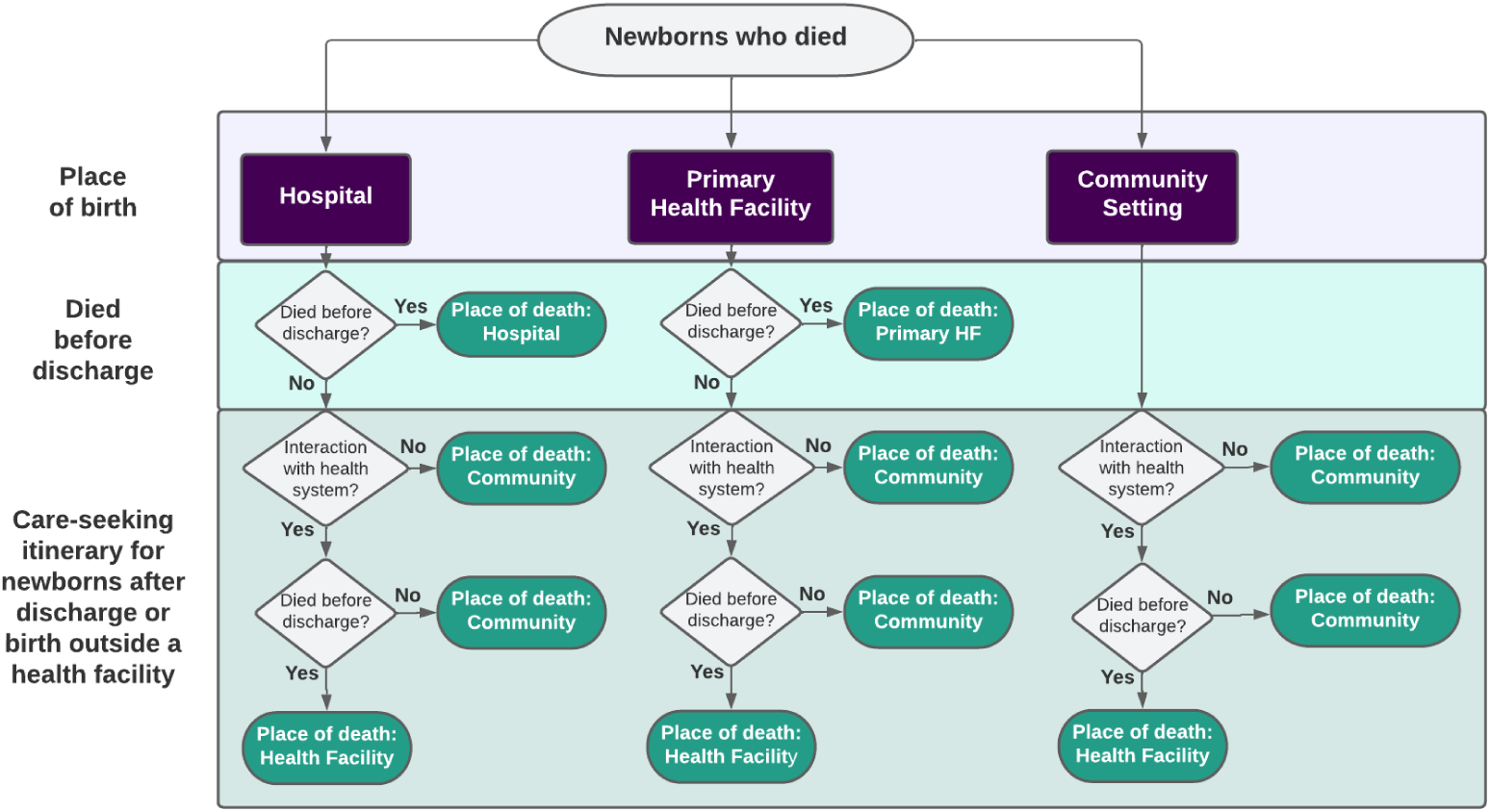
Care-seeking itinerary from birth to death for newborns who died in Malawi, Mozambique, and Tanzania.

For caretakers of newborns who died, we calculated the proportion who reported having concerns related to delays 1 and 2 in the Three Delays Model that affected their decision and ability to seek care for their newborn’s fatal illness.

#### Facility readiness to manage small and sick newborn care

We used the work of Moxon et. al (18,46,47), WHO guidance (8), and selected HFA questionnaires (23–25) to develop a measure of facility readiness to manage care for SSNs. Due to limited data availability, we were unable to include all SSN care interventions identified in the literature in this measure. Ultimately, we included seven interventions required to care for SSNs: essential newborn care (ENC)^2^, breastfeeding, resuscitation, prevention of mother-to-child transmission of HIV (PMTCT), kangaroo mother care (KMC), antibiotics for infection, and a cross-cutting domain for general readiness items. Detailed definitions of items included for each intervention and differences in data availability by item and country are available in the supplementary material (**Table A-2**). Because most of these items were collected in the labor and delivery module of the HFA questionnaire, we limited our analysis to facilities offering labor, delivery and newborn services (55% of facilities in Malawi, 85% in Mozambique, 80% in Tanzania) (see supplementary material **Table A-1** for final sample sizes).

For each facility included in the analysis, we defined binary (0/1) indicators^3^ for each item indicating whether the item was available and functional/non-expired at the time of the HFA. For each HFA, we calculated intervention-specific readiness scores for each of the six intervention domains and the cross-cutting domain as an unweighted average of available items in that domain. We then calculated an overall SSN care readiness score for each HFA by averaging the domain scores. We then disaggregated overall SSN readiness scores by health facility type. All analyses accounted for the HFA survey design (stratification and survey weights), as applicable.

#### Readiness-adjusted coverage of institutional deliveries for SSN care

To calculate readiness-adjusted institutional delivery coverage for small and sick newborn care in each country, we linked data on place of birth from the most recent HH survey (2015 for all countries) to the overall and intervention-specific SSN care readiness scores calculated from HFA data. HFAs for this analysis were selected based on the time period of HFA data collection overlapping with the reference period for the place of birth question in the country’s HH survey. For Mozambique, because no HFA met this criterion, the 2018 SARA was used.

We used an ecological approach, as validated in previous studies (48–50), to link each birth in the HH datasets to stratum-specific intervention-specific and overall readiness scores calculated from HFAs. First, for each country, we mapped response options for place of birth in the HH survey onto health facility types from the HFAs. We then assigned each birth in the HH dataset to a stratum, where stratum was defined as the place of birth (health facility type or home) and administrative area (district in Malawi, province in Mozambique, region in Tanzania). Based on the assigned stratum, each birth was linked to a SSN care readiness score, calculated as the stratum average readiness score from the corresponding HFA. For example, a baby who was born at a public health center in Mwanza region, Tanzania, would have been assigned the average readiness score for all public health centers in Mwanza region included in the Tanzania 2014-15 SPA. For 30% (51/168) of administrative area-facility type pairs for Malawi, 27% (9/33) for Mozambique, and 42% (151/360) for Tanzania, the national average readiness score for all facilities of a given type was used because there was no data collected in the HFA for any facility of the given type in the particular administrative area. Women who did not deliver in health facilities were assigned a readiness score of zero.

To estimate readiness-adjusted coverage of institutional deliveries in each country, we assigned each birth a value of zero or one (zero if the birth took place at home and one if the birth took place at a health facility of any type). We multiplied this value (0 or 1) by the readiness score (both overall and intervention-specific) for each birth. At a population level, this approach reduces the “institutional birth” coverage by the gap between measured and perfect facility readiness and accounts for differences in facility readiness by facility type. We calculated readiness-adjusted coverage at national level and by facility type by taking the mean of the products (readiness score multiplied by 0 or 1), accounting for both HH and HFA survey designs using the HH weights to generate point estimates and a jackknife approach to estimate the standard errors, where the standard error was derived from the distribution generated by withholding each HH cluster and each health facility (51). We also disaggregated this analysis by whether the baby died in the neonatal period; however, as we found no statistically significant differences between babies who died in the newborn period (days 0-27) and those who survived to day 28, we present these analyses for all live births.

### Statistical software

All analyses were completed using Stata/SE version 17.0 (52), R 4.0.3 (53), and R Studio (54).

## Findings

### National time trends in place of birth

Across all three countries, the proportion of babies born at home substantially decreased over time, as more births shifted to health facilities (**Figure *2***). The largest decrease in the proportion of births occurring at home was in Malawi, which experienced a 37% decrease in home births between 2000-2015, followed by Mozambique (22%) and Tanzania (17%) (***Figure 2–A,C,E***). In Malawi, this decrease occurred rapidly from 2006-2013, before leveling off with just 7% of births occurring at home by 2015. In Mozambique and Tanzania, comparatively, the decrease over time was relatively modest, resulting in 29% of births in Mozambique and 36% of births in Tanzania occurring at home by 2015. Breaking down the distribution of births by facility level and managing authority revealed that the decline in home births from 2000 to 2015 in Malawi and Tanzania was largely related to increases in births at public (government) primary health facilities (29% increase in Malawi and 8% in Tanzania), with smaller increases at public hospitals (9% increase in Malawi and 5% in Tanzania and virtually no change in the non-governmental and private sectors (**Figure 2–B,D,F**). In Mozambique, a larger increase was seen in the proportion of births at public hospitals (5% increase) compared to public primary facilities (1% increase), although similar to Malawi and Tanzania, the largest proportion of health facility births throughout the time period remained at public primary facilities. While there was also a large increase in the proportion of births at “Other public” health facilities in Mozambique from 2011 to 2015, this was an artifact of the categorization of health facilities in the 2015 AIS.

**Figure 2.**
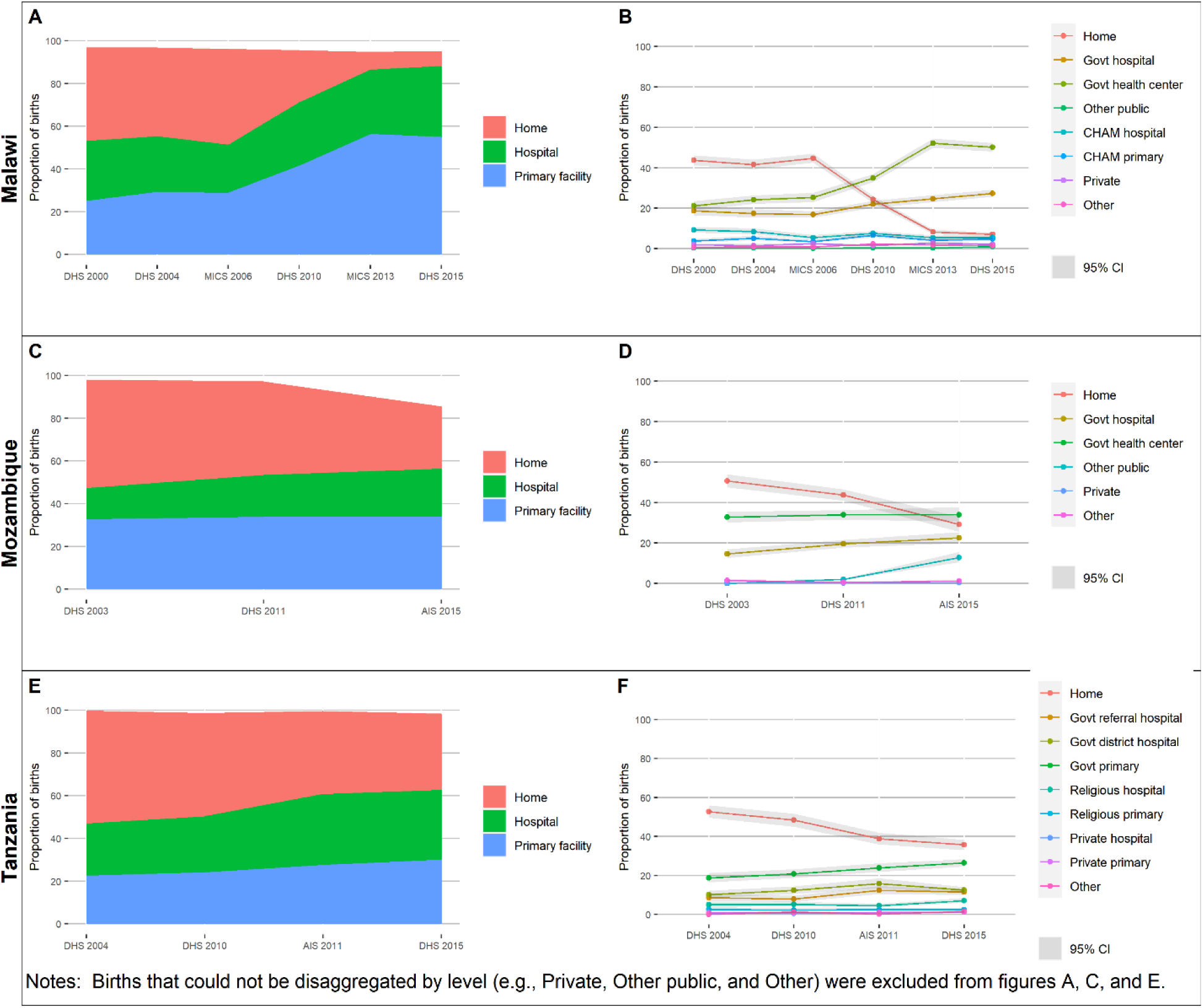
Proportion of births by place of birth, over time in Malawi, Mozambique, and Tanzania.

### Care-seeking behaviors for newborns who died

Exploring the care-seeking behaviors for newborns who died provides information on whether these newborns reached hospitals and/or primary health facilities around delivery, illness, and death. Across countries, at least half of the deceased newborns born in health facilities (hospital or primary) died before discharge (58% in Malawi, 46% in Mozambique, and 70% in Tanzania) (**Figure A-1**). The percentage of these deceased newborns who died at their birth facility is much higher for births at hospitals compared to primary facilities. While some deceased newborns were discharged after birth and died in the community without further interaction with the health system, many returned to a health facility (or followed a referral to another facility) after they were discharged from their birth facility. Most of the newborns who were discharged from their birth facilities did not develop their fatal illnesses until after discharge (94% in Malawi, 69% in Mozambique, 83% in Tanzania). Among deceased newborns who were born outside of health facilities (i.e., in a community setting or en route to a facility), the majority in all three countries had no interaction with the formal health system before death (56% in Malawi, 77% in Mozambique, and 57% in Tanzania). Another 3-4% of newborns in each country died on route to a health facility.

To better understand why care was not sought for many newborns’ fatal illnesses, we analyzed concerns related to delays in seeking facility-based care reported by caretakers of newborns who died. These concerns included a perceived lack of need, challenges in reaching health service delivery points, and quality of care (**Figure A-2**). Notably, in all three countries, over half of all these caretakers – 69% in Malawi, 59% in Mozambique, and 71% in Tanzania – did not report any concerns that deterred them from seeking care. Among caretakers who did express concerns, a wide range of concerns were identified. In Malawi, the most commonly reported concerns were that someone other than the caretaker was responsible for making this decision and that care-seeking took too much time away from the caretaker’s regular duties. In Tanzania, the most reported concerns included these same two concerns, in addition to concerns that the newborn was too sick to travel and would die no matter what. In Mozambique, a perception that the newborn was not sick enough to need health care and that health facilities were too far to travel were the most cited concerns. The mean number of concerns identified by caretakers with at least one concern was much higher for Malawi (11.96, sd=0.97) and Tanzania (12.29, sd=0.85) than for Mozambique (1.42, sd=0.63).^4^

### Facility readiness to manage small and sick newborns

Facility readiness scores provide insight into the third delay of the Three Delays Model – with lower scores indicating SSNs may face greater delays in accessing high-quality appropriate care *after* reaching a health facility due to inputs required for SSN health interventions not being available at the facility. An intervention-specific readiness score of 100 would indicate that, on average, health facilities in a given country had all items measured for that intervention available at the time of the HFA (see supplementary material **Table A-2** for a description of items used to calculate intervention-specific readiness scores for each country). No facilities in Malawi and Tanzania and only a small proportion of facilities in Mozambique (2.6%) were ready to deliver SSN care services (i.e., a readiness score of 100%). The overall SSN care facility readiness mean scores ranged from 53.7 out of 100 (95% CI: 52.6-54.7) in Tanzania to 65.1 (95% CI: 64.1-66.1) in Malawi, and 80.7 in Mozambique (95% CI: 80.1-81.3) (**Figure 3**). In Malawi, intervention-specific readiness was highest for resuscitation and general items (79.8 and 78.6, respectively) and lowest for KMC (33.7). In Mozambique, both breastfeeding and essential newborn care readiness scores were over 94, while resuscitation readiness was 60.1. Tanzania had the highest intervention-specific readiness score for general items (72.6), followed by breastfeeding (69.3) and resuscitation (65.0) while the lowest intervention-specific readiness score was for KMC (19.3).

**Figure 3.**
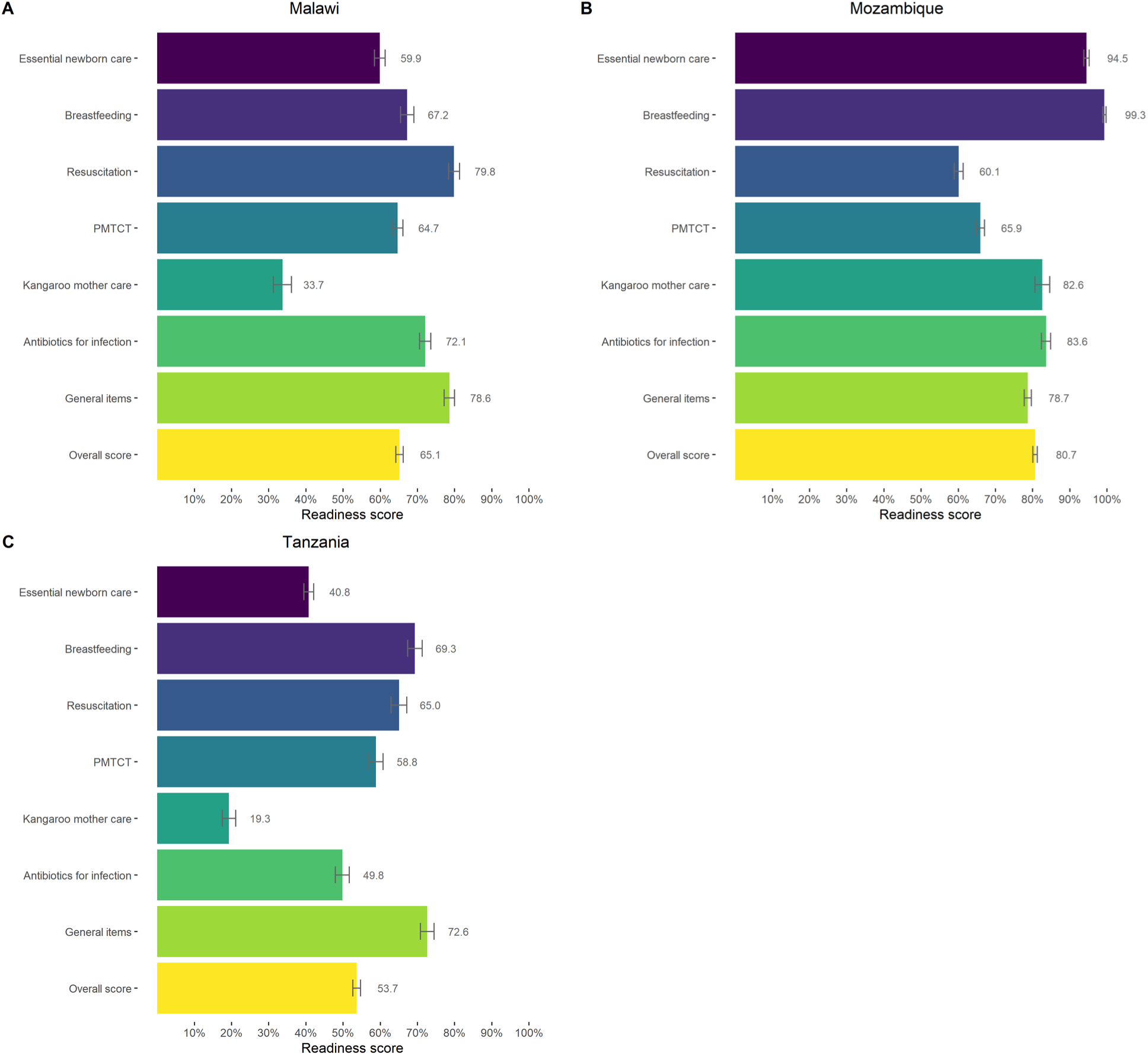
Overall and intervention-specific facility readiness scores to manage small and sick newborn care in Malawi, Mozambique, and Tanzania.

For all three countries, readiness varied by facility type, with hospitals generally more ready to care for small and sick newborns than primary health facilities (**Figure 4**). Across countries, within each managing authority category, facility readiness decreased as facility level decreased. In Malawi, average overall facility readiness among hospitals was 74 percentage points (pp), compared to 63 pp for lower-level, or primary, facilities (11 pp gap). In Mozambique, overall readiness of hospitals was 90 pp, compared to 80 pp in primary facilities (10 pp gap), and in Tanzania, overall readiness of hospitals was 69% compared to 55% among primary facilities (14 pp gap). In Tanzania, there were larger differences between referral and lower-level facilities, with a 21 pp gap between regional hospitals and dispensaries/clinics. The national hospitals in Tanzania had lower scores relative to regional hospitals driven by poor availability of items for KMC. Generally, facilities of the same level but different managing authorities performed similarly, although in Tanzania, overall SSN care readiness in private hospitals was more comparable to public primary health centers than to public referral hospitals. We could not disaggregate private facilities by facility level in Malawi and Mozambique due to survey limitations, but generally private facilities had lower readiness on average than other facility types. The wide variability in SSN care readiness in private facilities in Mozambique was a result of very few private facilities in the data set (3 out of 1,391 total facilities in the analysis).

**Figure 4.**
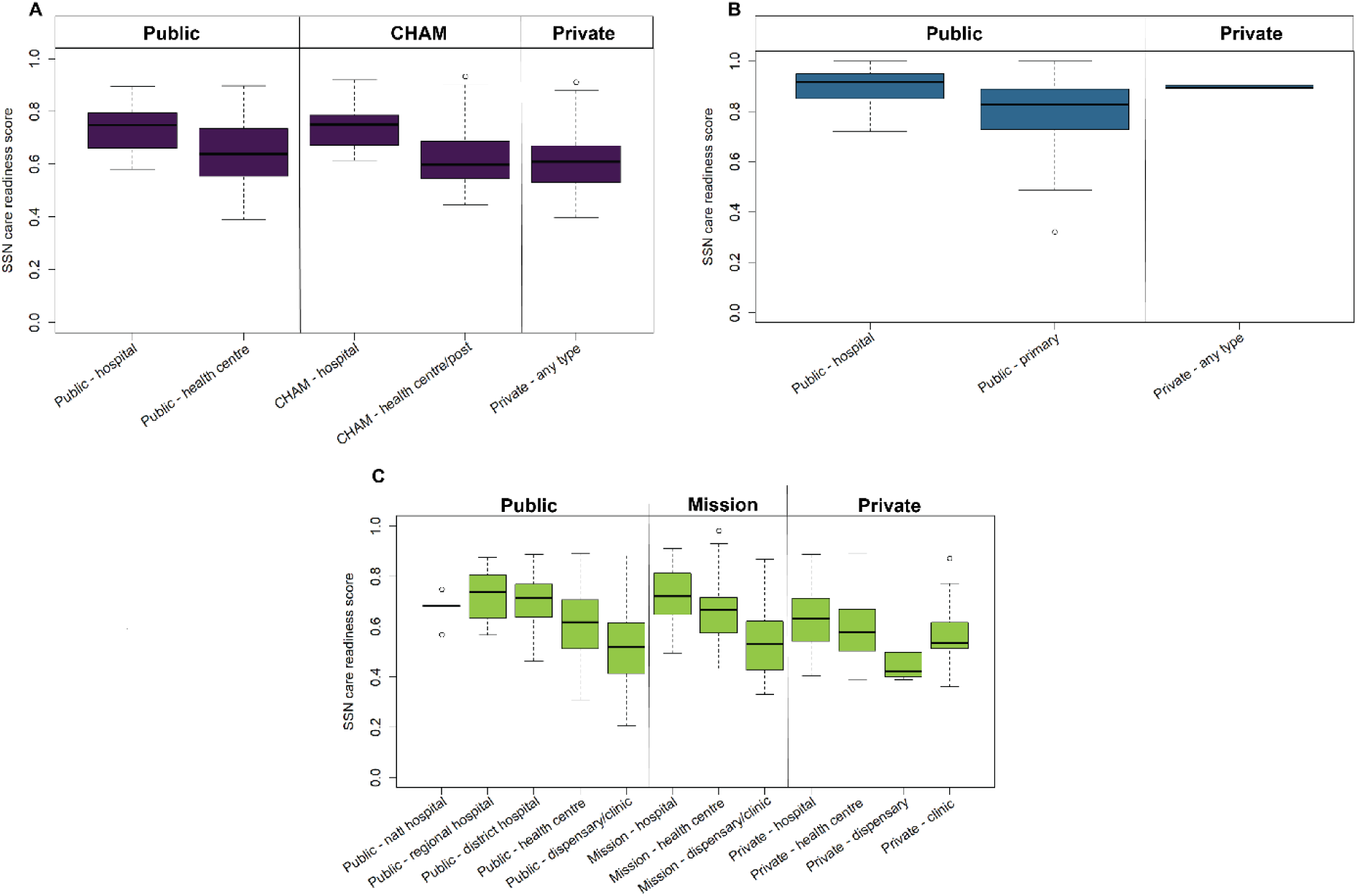
Overall readiness score to care for small and sick newborns by health facility type in Malawi, Mozambique, and Tanzania.

Figure 5 and **Table A-3** show the contribution of individual items to the SSN care readiness scores. In Malawi, across interventions, there was a low proportion of staff recently trained in neonatal care, including thermal care, cord care, newborn nutrition counseling, PMTCT, infection management, KMC and resuscitation (Figure 5-**A**). This trend was not limited to Malawi, with Mozambique and Tanzania showing similar gaps in availability of trained health workers (**Figure 5-B, C**). In all three countries, there were gaps in basic commodities, although the specific items varied by country and included waste receptacles in Malawi and Tanzania and soap and water for handwashing in Mozambique. There were large gaps in the availability of items such as a thermometer for low body temperature and Infant and Young Child Feeding (IYCF) counseling guidelines across countries. It is important to note that there is variation in the number of items included in interventions for each country driven by availability of data in the selected HFAs (**Table A-2**). The Mozambique HFA generally included fewer items for each intervention, whereas the Tanzania HFA included some additional items for interventions for which availability was particularly low (linen for drying the baby and Vitamin K).

**Figure 5.**
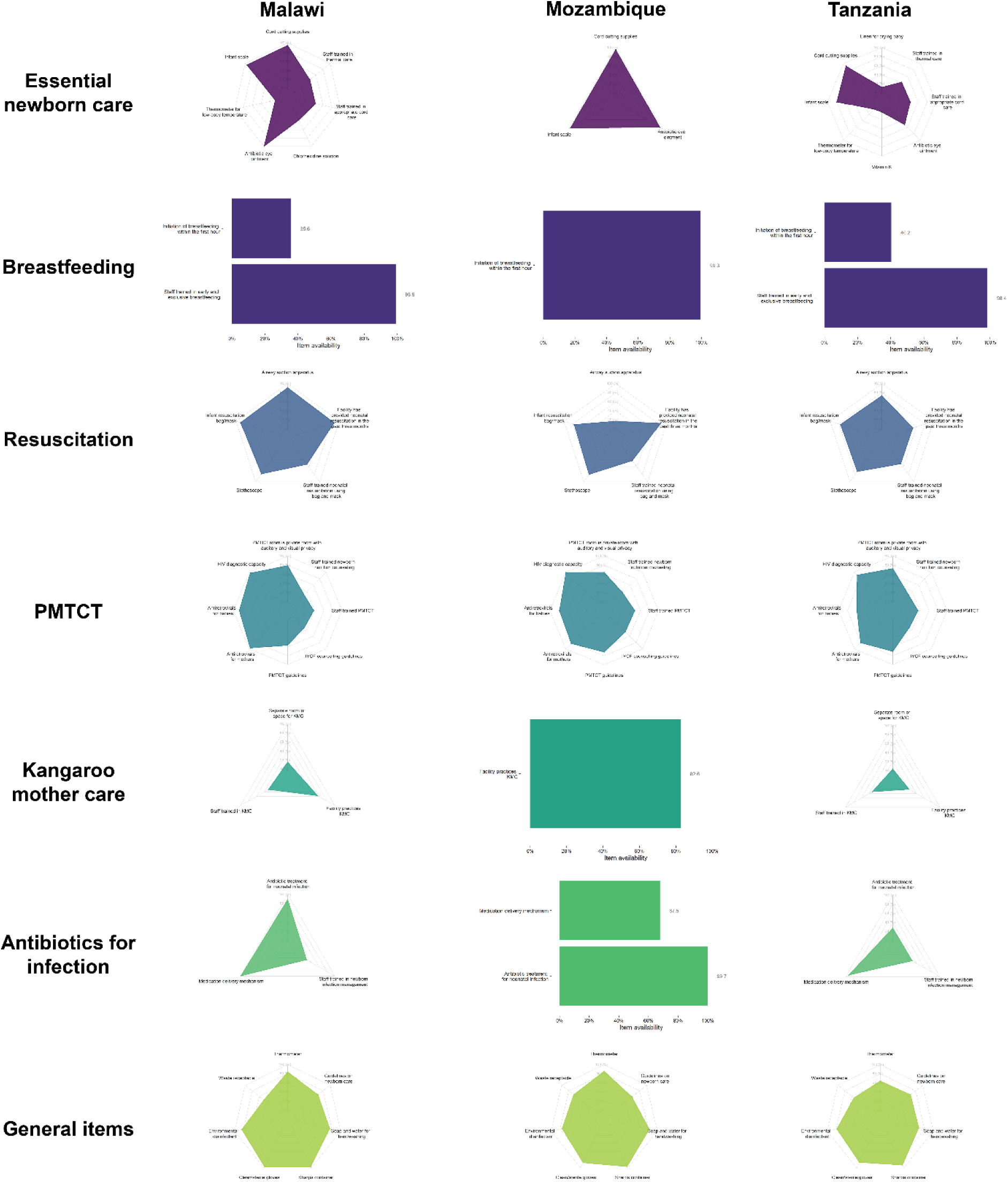
Availability of readiness items to manage small and sick newborns, by intervention, in Malawi, Mozambique, and Tanzania.

### Readiness-adjusted coverage of institutional delivery

After adjusting for readiness to manage small and sick newborns, coverage of institutional delivery in all three countries declined (Figure 6). The largest gap between unadjusted and overall readiness-adjusted coverage was in Malawi, where the vast majority of births (91%, 95% CI: 90-92) took place in health facilities. After adjusting for overall facility readiness to manage SSN care, coverage dropped to 62% (95% CI: 60-63), a gap of 30 pp. In Malawi, most of the intervention-specific readiness-adjusted coverage levels were similar to the overall readiness-adjusted coverage level, with the exception of KMC (38%; 95% CI: 35-42). For Mozambique, overall SSN care readiness-adjusted institutional delivery coverage was 56% (95% CI: 49-66), 14 pp lower than the service contact coverage value (70%, 95% CI: 66-73). Intervention-specific readiness-adjusted coverage values were relatively variable in Mozambique, ranging from 44% (95% CI: 37-51) for PMTCT to 69% (95% CI: 59-80) for breastfeeding. For Tanzania, overall SSN care readiness-adjusted coverage was 40% (95% CI: 37-43), a gap of 24 pp compared to the service contact coverage value of 63% (95% CI: 60-65). Intervention-specific readiness-adjusted coverage was also variable in Tanzania, ranging from 25% (95% CI: 22-29) for KMC to 52% (95% CI: 48-56) for general items.

**Figure 6.**
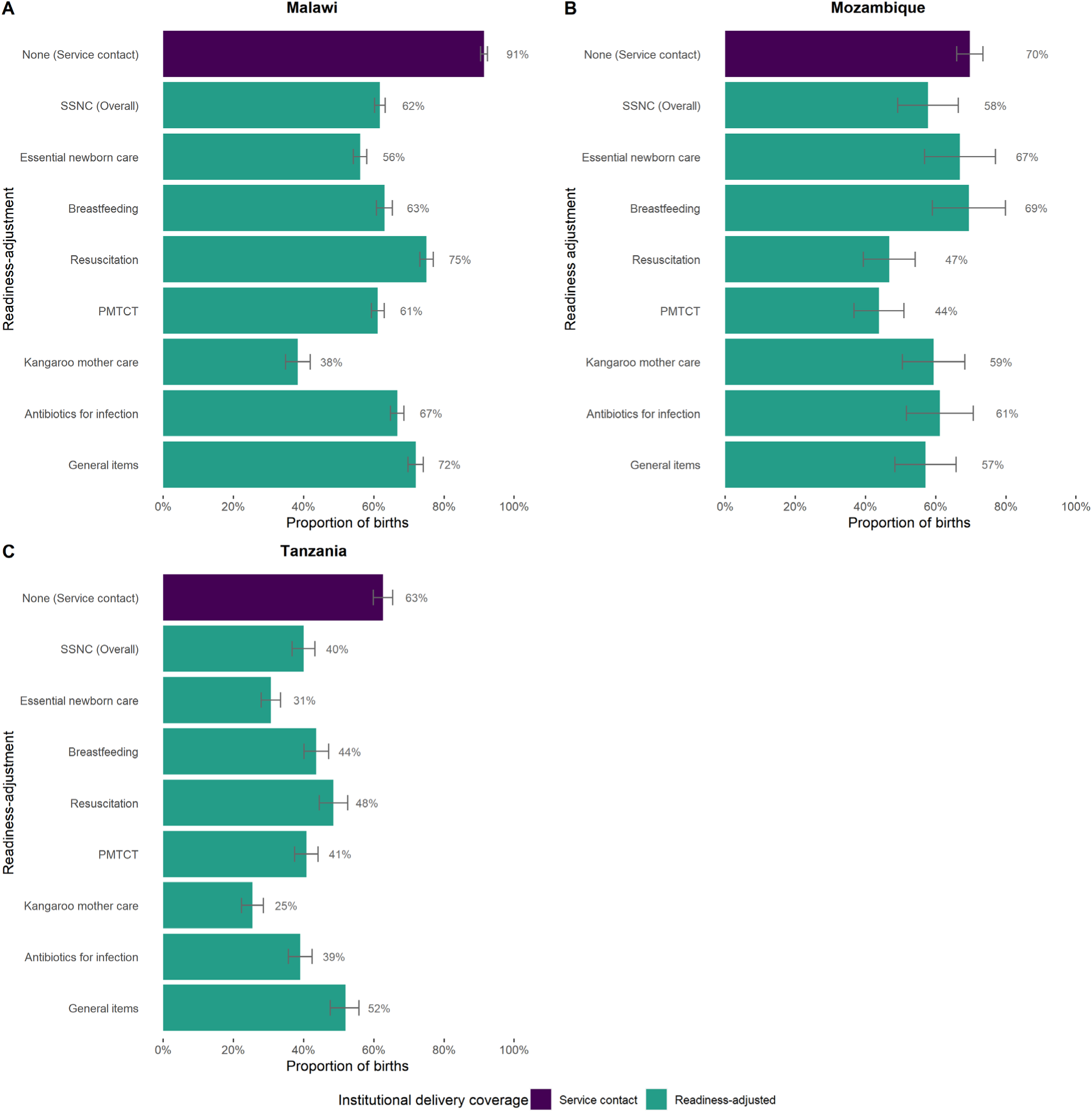
Service contact and small and sick newborn care readiness-adjusted institutional delivery coverage in Malawi, Mozambique, and Tanzania.

Comparing unadjusted and overall SSN care readiness-adjusted proportions of births categorized by birth location revealed gaps in effective coverage for all facility types in Malawi, Mozambique, and Tanzania (Figure 7). In Malawi, where most women gave birth in public primary health facilities, there was an 18 pp drop in coverage at public primary facilities and an 8 pp drop in coverage at public hospitals. In Mozambique, there was a small drop (2 pp) in coverage at hospitals, but a larger drop of 9 pp at primary facilities, where most facility births occurred. In Tanzania, there were significant gaps in readiness at all levels, including a 50% reduction in readiness-adjusted coverage at public dispensaries.

**Figure 7.**
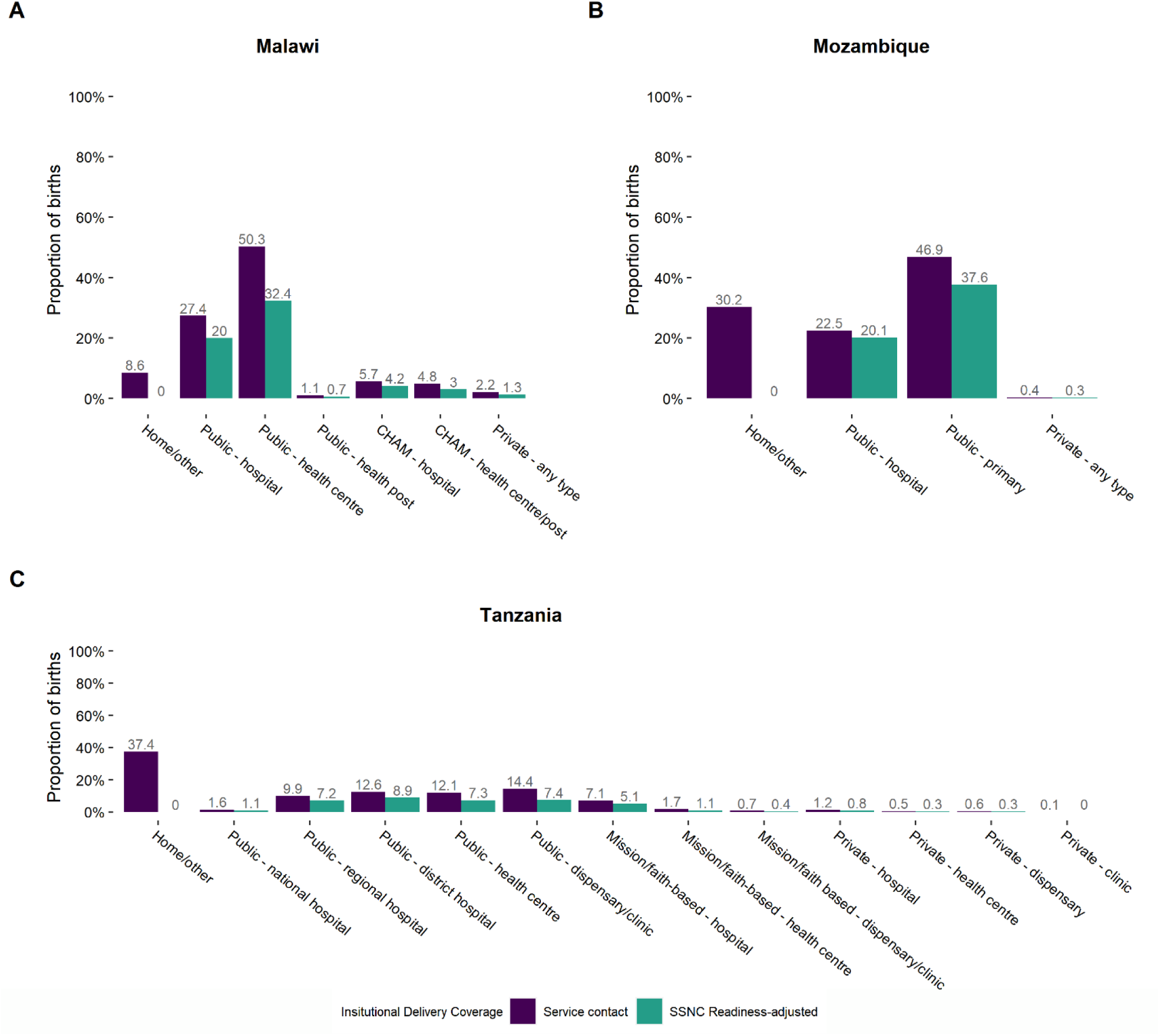
Service contact and small and sick newborn care readiness-adjusted place of birth distribution in Malawi, Mozambique, and Tanzania.

## Discussion

This study provides evidence that small and sick newborns in Malawi, Mozambique, and Tanzania may face substantial delays in access to high-quality care. We found that coverage of institutional delivery is substantially lower after adjusting for facility readiness to manage small and sick newborn care, with decreases of 30 pp in Malawi, 14 pp in Mozambique, and 24 pp in Tanzania. While delivery trends suggest more SSNs may reach facilities during the critical window of labor, delivery and the first week of life – overcoming the first and second delays in the Three Delays Model at the time of birth – these newborns may remain unable to access lifesaving interventions. Therefore, strategies aimed at addressing the delay in receipt of adequate and appropriate care after reaching a health facility (delay 3), particularly in the specific types of facilities where the largest proportion of newborns are born, would be particularly effective in these country contexts.

Our results support existing literature that show an increase in the proportion of births taking place in facilities over time in low- and middle-income countries (LMICs) (14,16). Multiple studies have explored individual-level and systems-level drivers of this trend (14–16,55,56), which include deliberate efforts by governments to increase access to facility-based delivery care (16,57). The impact of efforts to increase facility deliveries on neonatal mortality has been debated (58,59), and given low SSN care facility readiness at many facilities shown in this study, efforts to simply improve facility access at the time of birth in these countries are unlikely to be sufficient to substantially reduce neonatal mortality. While our results showed higher readiness among hospitals, many hospitals still lacked readiness items required for effective delivery of interventions for small and sick newborns. Thus, strategies that focus only on shifting deliveries to hospitals are also likely to be inadequate. Broader strategies, which include quality improvement initiatives at facilities, must also be considered. These strategies should target the facilities where newborns are born, such as first-level government facilities that offer delivery services. Our analysis corroborates findings by Wang, et al (60) that the increase in facility deliveries in Malawi, Mozambique, and Tanzania is largely attributed to increases in deliveries at public first level facilities.

Key areas for quality improvement of health facilities identified in our analysis include ensuring availability of basic commodities for newborns and sustaining a health workforce adequately trained in neonatal interventions. Since ENAP’s conception, stakeholders have identified limited health worker numbers and capacity as a key barrier to implementation of newborn care interventions (18,44,61–65). Countries and partners have employed a variety of strategies to address this limitation (66), but recent studies show that providers in LMICs still have relatively poor knowledge of newborn care interventions (67,68). Variation in intervention-specific readiness scores within the three countries in our analysis suggests that different interventions may need to be prioritized for each country to maximize the impact on neonatal survival. These interventions should be considered in both public and private health facilities, as our results showed facilities of the same level, regardless of managing authority, had similar readiness to deliver SSN care, contradicting a common perception that private health facilities offer better care than public facilities. In one exception, private hospitals in Tanzania had lower readiness to deliver SSN care compared to public referral hospitals. Additionally, as large proportions of newborns in Mozambique (29%) and Tanzania (36%) continue to be born in the community, efforts to improve the quality of care at facilities will only be most effective when combined with interventions targeting the first and second delays to enable newborns to reach services at health facilities.

Our analysis of care-seeking behaviors for newborns who died in Malawi, Mozambique, and Tanzania showed that many of these newborns who were born in facilities died before discharge, likewise suggesting the facilities in which they were born may not have been adequately prepared to manage their care (delay 3). While a much larger percentage of newborns born at hospitals died at their place of birth, compared to newborns born at primary facilities, this likely reflects birth complications or other difficult conditions which may be associated with seeking out delivery care from a hospital. Other studies have explored this relationship and have shown that delays pregnant women face reaching the most capable delivery facility may contribute to mortality of their newborns, especially if there are complications (69,70). In this study, we centered the newborn experience after birth, while we recognize delays in care-seeking faced by the newborn’s mother during pregnancy, labor, and delivery before birth may ultimately impact newborn health outcomes. This analysis also revealed that a substantial number of newborns who died never reached health facilities for care, and others were discharged from a health facility after birth but ultimately died at home without further care-seeking. Some of these deaths may be attributed to delays in the decision to seek care (delay 1) and ability to access an appropriate source of care, including inadequately functioning referral systems (delay 2). Across countries, caretakers reported similar concerns around accessing health services to those identified by other studies in LMIC contexts, such as distance to the health facility; costs associated with care-seeking, including opportunity costs from time away from regular duties; and no one available to accompany the caretaker (44,71–75). Many respondents in Malawi and Tanzania also noted they did not have a need to seek care, as they thought traditional care was more appropriate. Other studies using social autopsy data have also reported a preference for traditional care (and the need to administer traditional and allopathic care sequentially as opposed to simultaneously) as a barrier to care-seeking (72,76). Given that over half of all caretakers in our datasets reported no concerns related to care-seeking for their newborn’s fatal illness, influencers on decision-making around newborn care-seeking should be further explored.

While this analysis provided useful insights into access to care for small and sick newborns in Malawi, Mozambique, and Tanzania, we noted several limitations. Because this study relied on available survey data, the timeliness of survey data, lack of standardization of surveys across time and countries, small sample sizes (VASA), and data availability (HFAs) were limitations. Limited availability of data on items required for newborn care interventions with significant variation by country HFA – which limited comparability of readiness scores between countries in our analysis – and lack of consensus on guidelines to define interventions required for SSN care have been previously documented by Moxon and colleagues (46,47). In their analysis of the most used service availability assessment tools (including SPA and SARA), they found the neonatal interventions best represented in these tools are those promoted via vertical programming, including neonatal resuscitation, ENC, and PMTCT. The focus of SPA and SARA on items available in lower-level facilities, and correspondingly our analysis on first-level interventions for SSNs expected to be available in these facilities, is likely to mask some of the readiness differential between hospitals, which provide secondary- and tertiary-level SSNC interventions, and primary facilities and may lead to an underestimate of readiness-adjusted coverage in countries with high proportions of hospital deliveries. However, given that none of the countries in this analysis had large proportions of hospital deliveries, the effect of this bias on the overall readiness-adjusted estimates would likely have been minimal. We were also not able to capture components of effective referral and transportation.

In addition, we were unable to measure the full impact each of the three delays may have on access to care for SSNs in Malawi, Mozambique, and Tanzania. Since we did not have survey data that captured our target population of SSNs, we used all births as our population of interest, which may mask care-seeking differences between SSNs and other newborns. Likewise, without data on the proportion of newborns who needed each SSN service, we used institutional delivery as our service contact coverage measure. Among newborns who were born in health facilities, discharged after birth, and died at home without further care-seeking, most became sick after discharge and likely delayed care-seeking (delay 1 or 2). However, among the newborns who were sick before discharge, it is unclear if the facility was unable to provide adequate care (delay 3) or if the illness worsened upon returning home and further care-seeking was delayed (delays 1 or 2). Likewise, for newborns who were born in health facilities, discharged, and ultimately returned to a health facility where they died, it is unknown if they delayed care-seeking (delay 1 or 2), if the facility did not provide adequate care (delay 3), or both. For the third delay, we could not measure the full effective coverage cascade as proposed by Amouzou et. al (19) and the WHO Working Group (20), and while readiness is a necessary input to high-quality care, it does not indicate if newborns receive high-quality services. Due to data limitations, our analysis stopped at readiness-adjusted coverage. Although this is not the final endpoint of the effective coverage cascade, it does provide valuable information on SSN care readiness while information on intervention delivery and quality at the service delivery level remains limited in LMICs. While our findings must be interpreted in light of these limitations, this study provides useful insights into the extent to which each of the delays of the Three Delays Model may impact access to care for SSNs in Malawi, Mozambique, and Tanzania.

## Conclusions

This analysis suggests that if we are to achieve the vision set out in ENAP and meet SDG 3 targets by 2030, substantial investment must be made to overcome delays in access to care for the most vulnerable newborns – those who are born small or sick. Addressing limited access to high-quality care for SSNs, who account for a disproportionate fraction of total neonatal mortality, is essential to drive down neonatal mortality rates by the end of the decade. As more women and newborns have access to health services in LMICs, ensuring life-saving interventions for small and sick newborns are available at the locations where newborns are born and seek care after birth is critical. Future research to address data gaps in HFAs, provide additional information on care-seeking behaviors, and assess facility readiness and quality to provide SSN care in particular geographic areas would help inform the development and prioritization of locally tailored interventions to improve neonatal survival.

## List of abbreviations

AIS: AIDS Indicator Survey
BEmOC: Basic emergency obstetric care
CEmOC: Comprehensive emergency obstetric care
CHW: Community health worker
DHS: Demographic and Health Survey
ENAP: Every Newborn Action Plan
ENC: Essential newborn care
HH: Household survey
HFA: Health facility assessment
IMPAC: Integrated Management of Pregnancy and Childbirth
IYCF: Infant and Young Child Feeding
KMC: Kangaroo mother care
LMIC: Low- or middle-income country
MICS: Multiple Indicator Cluster Survey
NMR: Neonatal mortality rate
PMTCT: Prevention of mother-to-child transmission of HIV
SARA: Service Availability and Readiness Assessment
SDG: Sustainable Development Goal
SPA: Service Provision Assessment
UN: United Nations
UNICEF: United Nations Children’s Fund
VASA: Verbal and social autopsy
WHA: World Health Assembly
WHO: World Health Organization

## Declarations

## Acknowledgements

The authors would like to acknowledge the Bill & Melinda Gates Foundation for supporting this project.

## Ethics approval

This is a secondary analysis and as such did not involve human subjects research.

## Authors’ contributions

LN, AS, and MKM contributed to conceptualizing the paper and analysis. LN and AS conducted the analysis and prepared the manuscript. All authors critically reviewed and revised the manuscript and approved the final manuscript.

## Availability of data and materials

HH survey data are available in public, open access repositories (DHS and AIS via the DHS Program website, and MICS via the UNICEF website). HFA data for Malawi and Tanzania SPAs are also available in public, open access repositories through the DHS Program. HFA data for the Mozambique 2018 SARA can be requested by submitting <u>this Google Form</u> and then emailing. De-identified VASA data subsets used in this analysis for each country can be accessed <u>here</u>.

## Footnotes

1 The Malawi MICS 2006 and Malawi MICS 2013-14 only collected information on births within the two years preceding the surveys. The Mozambique AIS 2015 only collected information on births within 32 months preceding the survey.

2 Essential newborn care is a package of interventions expected to be provided to every newborn at the time of delivery and includes drying, hygienic cord care, vitamin K, and eye care.

3 Binary indicators were created for all readiness items except for items derived from health provider sections of the HFAs. These indicators are instead defined as the proportion of health providers providing delivery or newborn care services that have been trained in or carried out a particular intervention within a specified time period.

4 Differences in cta collection methods between CHERG VASA (Malawi and Tanzania) and COMSA VASA (Mozambique). Data on concerns was collected by the data collector marking all applicable concerns from a checklist based on an open-ended response from the respondent.

## References

1. United Nations Inter-agency Group for Child Mortality Estimation [Internet]. 2021 [cited 2022 Feb 24]. Available from: https://childmortality.org/data

2. United Nations Inter-Agency Group for Child Mortality Estimation. Levels and trends in child mortality [Internet]. United Nations Children’s Fund (UNICEF); 2021 Dec [cited 2022 Feb 24]. Available from: https://data.unicef.org/resources/levels-and-trends-in-child-mortality/

3. World Health Organization. Every Newborn: an action plan to end preventable deaths [Internet]. WHO, UNICEF, EWEC; 2014 [cited 2022 Feb 24]. Available from: https://www.who.int/publications-detail-redirect/9789241507448

4. World Health Organization. Standards for improving the quality of care for small and sick newborns in health facilities [Internet]. World Health Organization; 2020 [cited 2022 Jan 11]. v, 143 p. Available from: https://apps.who.int/iris/handle/10665/334126

5. Lawn JE, Blencowe H, Oza S, You D, Lee AC, Waiswa P, et al. Every Newborn: progress, priorities, and potential beyond survival. The Lancet. 2014 Jul;384(9938):189–205.

6. Lee AC, Kozuki N, Cousens S, Stevens GA, Blencowe H, Silveira MF, et al. Estimates of burden and consequences of infants born small for gestational age in low and middle income countries with INTERGROWTH-21st standard: analysis of CHERG datasets. BMJ. 2017 Aug 17;358:j3677.

7. Katz J, Lee AC, Kozuki N, Lawn JE, Cousens S, Blencowe H, et al. Mortality risk in preterm and small-for-gestational-age infants in low-income and middle-income countries: a pooled country analysis. The Lancet. 2013 Aug 3;382(9890):417–25.

8. World Health Organization. Survive and thrive: transforming care for every small and sick newborn [Internet]. World Health Organization; 2019 [cited 2022 Feb 24]. x, 150 p. Available from: https://apps.who.int/iris/handle/10665/326495

9. Bhutta ZA, Das JK, Bahl R, Lawn JE, Salam RA, Paul VK, et al. Can available interventions end preventable deaths in mothers, newborn babies, and stillbirths, and at what cost? The Lancet. 2014 Jul 26;384(9940):347–70.

10. Lassi ZS, Middleton PF, Crowther C, Bhutta ZA. Interventions to Improve Neonatal Health and Later Survival: An Overview of Systematic Reviews. EBioMedicine. 2015 Aug 1;2(8):985–1000.

11. Lawn JE, Mwansa-Kambafwile J, Horta BL, Barros FC, Cousens S. “Kangaroo mother care” to prevent neonatal deaths due to preterm birth complications. Int J Epidemiol. 2010 Apr;39 Suppl 1:i144–154.

12. Lee ACC, Cousens S, Wall SN, Niermeyer S, Darmstadt GL, Carlo WA, et al. Neonatal resuscitation and immediate newborn assessment and stimulation for the prevention of neonatal deaths: a systematic review, meta-analysis and Delphi estimation of mortality effect. BMC Public Health. 2011 Apr 13;11 Suppl 3:S12.

13. Thaddeus S, Maine D. Too far to walk: Maternal mortality in context. Soc Sci Med. 1994 Apr 1;38(8):1091–110.

14. Hill Z, Amare Y, Scheelbeek P, Schellenberg J. ‘ People have started to deliver in the facility these days ’ : a qualitative exploration of factors affecting facility delivery in Ethiopia. BMJ Open. 2019 Jun 1;9:e025516.

15. Diamond-Smith N, Sudhinaraset M. Drivers of facility deliveries in Africa and Asia: regional analyses using the demographic and health surveys. Reprod Health. 2015 Jan 16;12(1):6.

16. Montagu D, Sudhinaraset M, Diamond-Smith N, Campbell O, Gabrysch S, Freedman L, et al. Where women go to deliver: understanding the changing landscape of childbirth in Africa and Asia. Health Policy Plan. 2017 Oct 1;32(8):1146–52.

17. Goudar SS, Goco N, Somannavar MS, Kavi A, Vernekar SS, Tshefu A, et al. Institutional deliveries and stillbirth and neonatal mortality in the Global Network’s Maternal and Newborn Health Registry. Reprod Health. 2020 Dec 17;17(3):179.

18. Moxon SG, Lawn JE, Dickson KE, Simen-Kapeu A, Gupta G, Deorari A, et al. Inpatient care of small and sick newborns: a multi-country analysis of health system bottlenecks and potential solutions. BMC Pregnancy Childbirth. 2015 Sep 11;15(2):S7.

19. Amouzou A, Leslie HH, Ram M, Fox M, Jiwani SS, Requejo J, et al. Advances in the measurement of coverage for RMNCH and nutrition: from contact to effective coverage. BMJ Glob Health. 2019 May 1;4(Suppl 4):e001297.

20. Marsh AD, Muzigaba M, Diaz T, Requejo J, Jackson D, Chou D, et al. Effective coverage measurement in maternal, newborn, child, and adolescent health and nutrition: progress, future prospects, and implications for quality health systems. Lancet Glob Health. 2020 May;8(5):e730–6.

21. ICF. The DHS Program website. Funded by USAID. [cited 2022 Jan 5]. Available Datasets. Available from: http://www.dhsprogram.com

22. UNICEF. Surveys - UNICEF MICS [Internet]. [cited 2022 Jan 5]. Available from: https://mics.unicef.org/surveys

23. MoH/Malawi M of H, International ICF. Malawi Service Provision Assessment 2013-14. 2014 Nov 1 [cited 2022 Apr 10]; Available from: https://dhsprogram.com/publications/publication-spa20-spa-final-reports.cfm

24. Ministério da Saúde (MISAU), Instituto Nacional de Saúde (INS), Governo do Canadá, Organização Mundial da Saúde (OMS). Moçambique Service Availability and Readiness Assessment 2018: Inventário Nacional. MISAU/Moçambique, INS/Moçambique, Governo do Canadá, and OMS; 2020 Feb.

25. Welfare/Tanzania M of H and S, Health/Zanzibar M of, Statistics/Tanzania NB of, Statistician/Tanzania O of CG, International ICF. Tanzania Service Provision Assessment Survey 2014-2015. 2016 Feb 1 [cited 2022 Apr 10]; Available from: https://dhsprogram.com/publications/publication-spa22-spa-final-reports.cfm

26. National Statistical Office/Malawi and ORC Macro. Malawi Demographic and Health Survey 2000 [Internet]. Zomba, Malawi: National Statistical Office/Malawi and ORC Macro; 2001 Jun [cited 2022 Apr 12]. Available from: https://dhsprogram.com/publications/publication-fr123-dhs-final-reports.cfm

27. National Statistical Office - NSO/Malawi and ORC Macro. Malawi Demographic and Health Survey 2004 [Internet]. Calverton, Maryland: NSO/Malawi and ORC Macro; 2005 Dec [cited 2022 Apr 12]. Available from: https://dhsprogram.com/publications/publication-fr175-dhs-final-reports.cfm

28. National Statistical Office and UNICEF. Malawi Multiple Indicator Cluster Survey 2006, Final Report [Internet]. Lilongwe, Malawi: National Statistical Office and UNICEF; 2008 [cited 2022 Apr 12]. Available from: https://microdata.worldbank.org/index.php/catalog/1798

29. National Statistical Office - NSO/Malawi, ICF Macro. Malawi Demographic and Health Survey 2010 [Internet]. Zomba, Malawi: NSO/Malawi and ICF Macro; 2011 Sep [cited 2022 Apr 12]. Available from: https://dhsprogram.com/publications/publication-fr247-dhs-final-reports.cfm

30. National Statistical Office of Malawi. Malawi MDG Endline Survey 2014. Zomba, Malawi: National Statistical Office of Malawi; 2015 Jun.

31. National Statistical Office - NSO/Malawi and ICF. Malawi Demographic and Health Survey 2015-16 [Internet]. Zomba, Malawi: NSO and ICF; 2017 Feb [cited 2022 Apr 12]. Available from: https://dhsprogram.com/publications/publication-FR319-DHS-Final-Reports.cfm

32. Instituto Nacional de Estatística/Moçambique, Ministério da Saúde/Moçambique, and MEASURE DHS+/ORC Macro. Moçambique Inquérito Demográfico e de Saúde 2003 [Internet]. Calverton, Maryland, USA: Instituto Nacional de Estatística/Moçambique, Ministério da Saúde/Moçambique, and MEASURE DHS+/ORC Macro; 2005 Jan [cited 2022 Apr 12]. Available from: https://dhsprogram.com/publications/publication-FR161-DHS-Final-Reports.cfm

33. Ministerio da Saude - MISAU/Moçambique, Instituto Nacional de Estatística - INE/Moçambique and ICF International. Moçambique Inquérito Demográfico e de Saúde 2011 [Internet]. Calverton, Maryland, USA: MISA/Moçambique, INE/Moçambique and ICF International; 2013 Mar [cited 2022 Apr 12]. Available from: https://dhsprogram.com/publications/publication-FR266-DHS-Final-Reports.cfm

34. MISAU, Instituto Nacional de Estatística - INE, and ICF. Inquérito de Indicadores de Imunização, Malária e HIV/SIDA em Moçambique (IMASIDA) 2015 [Internet]. MISAU/Moçambique, INE, and ICF; 2018 Feb [cited 2022 Apr 12] p. Maputo, Moçambique. Available from: https://dhsprogram.com/publications/publication-AIS12-AIS-Final-Reports.cfm

35. National Bureau of Statistics - NBS/Tanzania and ORC Macro. Tanzania Demographic and Health Survey 2004-05 [Internet]. Dar es Salaam, Tanzania: NBS/Tanzania and ORC Macro; 2005 Dec [cited 2022 Apr 12]. Available from: https://dhsprogram.com/publications/publication-fr173-dhs-final-reports.cfm

36. National Bureau of Statistics - NBS/Tanzania and ICF Macro. Tanzania Demographic and Health Survey 2010 [Internet]. Dar es Salaam, Tanzania: NBS/Tanzania and ICF Macro; 2011 Apr [cited 2022 Apr 12]. Available from: https://dhsprogram.com/publications/publication-fr243-dhs-final-reports.cfm

37. Tanzania Commission for AIDS - TACAIDS, Zanzibar AIDS Commission - ZAC/Tanzania, National Bureau of Statistics - NBS/Tanzania, Office of the Chief Government Statistician - OCGS/Tanzania, ICF International. Tanzania HIV/AIDS and Malaria Indicator Survey 2011-12 [Internet]. Dar es Salaam, Tanzania: TACAIDS/Tanzania, ZAC/Tanzania, NBS/Tanzania, OCGS/Tanzania, and ICF International; 2013 Mar [cited 2022 Apr 12]. Available from: https://dhsprogram.com/publications/publication-ais11-ais-final-reports.cfm

38. Ministry of Health, Community Development, Gender, Elderly and Children - MoHCDGEC/Tanzania Mainland, Ministry of Health - MoH/Zanzibar, National Bureau of Statistics - NBS/Tanzania, Office of Chief Government Statistician - OCGS/Zanzibar, ICF. Tanzania Demographic and Health Survey and Malaria Indicator Survey 2015-2016 [Internet]. Dar es Salaam, Tanzania: MoHCDGEC, MoH, NBS, OCGS, and ICF; 2016 Dec [cited 2022 Apr 12]. Available from: https://dhsprogram.com/publications/publication-FR321-DHS-Final-Reports.cfm

39. Koffi AK, Mleme T, Nsona H, Banda B, Amouzou A, Kalter HD. Social autopsy of neonatal mortality suggests needed improvements in maternal and neonatal interventions in Balaka and Salima districts of Malawi. J Glob Health. 2015 Jun 17;5(1):010416.

40. Koffi AK, Kalter HD, Kamwe MA, Black RE. Verbal/social autopsy analysis of causes and determinants of under-5 mortality in Tanzania from 2010 to 2016. J Glob Health. 2020 Dec;10(2):020901.

41. Adriano A, Amouzou A, Aveika A, Black RE, Datta A, Duarte L, et al. Countrywide Mortaity Surveillance for Action (COMSA) - Mozambique: Mortality and Cause of Death in 2019.

42. Institute for International Programs. Johns Hopkins Bloomberg School of Public Health. [cited 2022 Apr 11]. Child Health Epidemiology Reference Group (CHERG). Available from: https://www.jhsph.edu/research/centers-and-institutes/institute-for-international-programs/completed-projects/child-health-epidemiology-reference-group/

43. Waiswa P, Kallander K, Peterson S, Tomson G, Pariyo GW. Using the three delays model to understand why newborn babies die in eastern Uganda. Trop Med Int Health. 2010;15(8):964–72.

44. Upadhyay RP, Rai SK, Krishnan A. Using Three Delays Model to Understand the Social Factors Responsible for Neonatal Deaths in Rural Haryana, India. J Trop Pediatr. 2013 Apr 1;59(2):100–5.

45. Applying the Three Delays Model: Improving access to care for newborns with danger signs [Internet]. Save the Children; 2013 Apr. Available from: https://www.healthynewbornnetwork.org/hnn-content/uploads/Applying-the-three-delays-model_Final.pdf

46. Moxon SG, Guenther T, Gabrysch S, Enweronu-Laryea C, Ram PK, Niermeyer S, et al. Service readiness for inpatient care of small and sick newborns: what do we need and what can we measure now? J Glob Health. 2018;8(1):010702.

47. Moxon SG, Blencowe H, Bailey P, Bradley J, Day LT, Ram PK, et al. Categorising interventions to levels of inpatient care for small and sick newborns: Findings from a global survey. PLOS ONE. 2019 Jul 11;14(7):e0218748.

48. Munos MK, Maiga A, Do M, Sika GL, Carter ED, Mosso R, et al. Linking household survey and health facility data for effective coverage measures: A comparison of ecological and individual linking methods using the Multiple Indicator Cluster Survey in Côte d’Ivoire. J Glob Health. 2018;8(2):020803.

49. Carter ED, Ndhlovu M, Eisele TP, Nkhama E, Katz J, Munos M. Evaluation of methods for linking household and health care provider data to estimate effective coverage of management of child illness: results of a pilot study in Southern Province, Zambia. J Glob Health. 2018;8(1):010607.

50. Carter ED, Leslie HH, Marchant T, Amouzou A, Munos MK. Methodological considerations for linking household and healthcare provider data for estimating effective coverage: a systematic review. BMJ Open. 2021 Aug 1;11(8):e045704.

51. Kott P. The delete-a-group jackknife. J Off Stat. 2001 Dec 1;17(4):521–6.

52. Stata Statistical Software: Release 17. Special Edition. College Station, Texas, USA: StataCorp; 2019.

53. R Core Team. R: A language and environment for statistical computing [Internet]. Vienna, Austria: R Foundation for Statistical Computing; 2020. Available from: https://www.R-project.org/

54. R Studio Team. R Studio: Integrated Development Environment for R [Internet]. Boston, MA, USA: RStudio; 2020. Available from: http://www.rstudio.com/

55. Moyer CA, Mustafa A. Drivers and deterrents of facility delivery in sub-Saharan Africa: a systematic review. Reprod Health. 2013 Aug 20;10(1):40.

56. Gebremichael SG, Fenta SM. Determinants of institutional delivery in Sub-Saharan Africa: findings from Demographic and Health Survey (2013–2017) from nine countries. Trop Med Health. 2021 May 26;49:45.

57. Bucagu M, Kagubare JM, Basinga P, Ngabo F, Timmons BK, Lee AC. Impact of health systems strengthening on coverage of maternal health services in Rwanda, 2000–2010: a systematic review. Reprod Health Matters. 2012 Jun 1;20(39):50–61.

58. Tura G, Fantahun M, Worku A. The effect of health facility delivery on neonatal mortality: systematic review and meta-analysis. BMC Pregnancy Childbirth. 2013 Jan 22;13:18.

59. Gabrysch S, Nesbitt RC, Schoeps A, Hurt L, Soremekun S, Edmond K, et al. Does facility birth reduce maternal and perinatal mortality in Brong Ahafo, Ghana? A secondary analysis using data on 119 244 pregnancies from two cluster-randomised controlled trials. Lancet Glob Health. 2019 Aug 1;7(8):e1074–87.

60. Wang W, Mallick L, Allen C, Pullum T. Effective coverage of facility delivery in Bangladesh, Haiti, Malawi, Nepal, Senegal, and Tanzania. PLoS ONE. 2019 Jun 11;14(6):e0217853.

61. Murphy GAV, Gathara D, Abuya N, Mwachiro J, Ochola S, Ayisi R, et al. What capacity exists to provide essential inpatient care to small and sick newborns in a high mortality urban setting? - A cross-sectional study in Nairobi City County, Kenya. PLoS ONE. 2018 Apr 27;13(4):e0196585.

62. Dickson KE, Simen-Kapeu A, Kinney MV, Huicho L, Vesel L, Lackritz E, et al. Every Newborn: health-systems bottlenecks and strategies to accelerate scale-up in countries. The Lancet. 2014 Aug 2;384(9941):438–54.

63. Mason E, McDougall L, Lawn JE, Gupta A, Claeson M, Pillay Y, et al. From evidence to action to deliver a healthy start for the next generation. The Lancet. 2014 Aug 2;384(9941):455–67.

64. Usman AK, Wolka E, Tadesse Y, Tariku A, Yeshidinber A, Teklu AM, et al. Health system readiness to support facilities for care of preterm, low birth weight, and sick newborns in Ethiopia: a qualitative assessment. BMC Health Serv Res. 2019 Nov 21;19(1):860.

65. Sami S, Amsalu R, Dimiti A, Jackson D, Kenyi S, Meyers J, et al. Understanding health systems to improve community and facility level newborn care among displaced populations in South Sudan: a mixed methods case study. BMC Pregnancy Childbirth. 2018 Aug 10;18:325.

66. Fung A, Hamilton E, Du Plessis E, Askin N, Avery L, Crockett M. Training programs to improve identification of sick newborns and care-seeking from a health facility in low- and middle-income countries: a scoping review. BMC Pregnancy Childbirth. 2021 Dec 14;21(1):831.

67. Murphy GAV, Gathara D, Mwaniki A, Nabea G, Mwachiro J, Abuya N, et al. Nursing knowledge of essential maternal and newborn care in a high-mortality urban African setting: A cross-sectional study. J Clin Nurs. 2019;28(5–6):882–93.

68. Arba A, Zana Z. Knowledge of Essential Newborn Care and Associated Factors among Nurses and Midwives: A Cross-Sectional Study at Public Health Facilities in Wolaita Zone, Southern Ethiopia, 2019. Int J Pediatr. 2020;2020:3647309.

69. Deviany PE, Setel PW, Kalter HD, Anggondowati T, Martini M, Nandiaty F, et al. Neonatal mortality in two districts in Indonesia: Findings from Neonatal Verbal and Social Autopsy (VASA). PloS One. 2022;17(3):e0265032.

70. Kalter HD, Setel PW, Deviany PE, Nugraheni SA, Sumarmi S, Weaver EH, et al. Modified Pathway to Survival highlights importance of rapid access to quality institutional delivery care to decrease neonatal mortality in Serang and Jember districts, Java, Indonesia. J Glob Health. 13:04020.

71. Nandan D, Misra SK, Jain M, Singh D, Verma M, Sethi V. Social Audits for Community Action: A tool to Initiate Community Action for Reducing Child Mortality. Indian J Community Med. 2005 Sep;30(3):78.

72. Bazzano AN, Kirkwood BR, Tawiah-Agyemang C, Owusu-Agyei S, Adongo PB. Beyond symptom recognition: care-seeking for ill newborns in rural Ghana. Trop Med Int Health. 2008;13(1):123–8.

73. Moyer CA, Johnson C, Kaselitz E, Aborigo R. Using social autopsy to understand maternal, newborn, and child mortality in low-resource settings: a systematic review of the literature. Glob Health Action. 2017 Dec 20;10(1):1413917.

74. Koffi AK, Libite PR, Moluh S, Wounang R, Kalter HD. Social autopsy study identifies determinants of neonatal mortality in Doume, Nguelemendouka and Abong-Mbang health districts, Eastern Region of Cameroon. J Glob Health. 2015 Jun;5(1):010413.

75. Koffi AK, Maina A, Yaroh AG, Habi O, Bensaïd K, Kalter HD. Social determinants of child mortality in Niger: Results from the 2012 National Verbal and Social Autopsy Study. J Glob Health. 2016 Jun;6(1):010603.

76. Nonyane BA, Kazmi N, Koffi AK, Begum N, Ahmed S, Baqui AH, et al. Factors associated with delay in care-seeking for fatal neonatal illness in the Sylhet district of Bangladesh: results from a verbal and social autopsy study. J Glob Health. 2016 Jun;6(1):010605.

