## Supplementary_Material for "Delays in accessing high-quality care for newborns in East Africa: An analysis of survey data in Malawi, Mozambique, and Tanzania"

**Table A- 1. Survey details.** Surveys used for analyses, including sample sizes and representativeness. Additional details on survey design and data collection procedures for household surveys and health facility assessments can be found in corresponding survey reports.

| Country | Survey | Sample size, survey | Sample size, analysis | Representativeness |
|---|---|---|---|---|
| **National time trends in place of birth** | | | | |
| Household surveys (HH) | | Interviewed women aged 15-49 | Live births[2] | Geographic representativeness |
| Malawi | DHS 2000 | 13,220 | 11,886 | National, regional, district |
| | DHS 2004 | 11,698 | 10,901 | National, regional, selected districts |
| | MICS 2006 | 26,259 | 10,288 | National, regional, district |
| | DHS 2010 | 23,020 | 19,880 | National, regional, district |
| | MICS 2013-14 | 24,230 | 7,494 | National, regional, district |
| | DHS 2015-16 | 24,562 | 17,148 | National, regional, district |
| Mozambique | DHS 2003 | 12,418 | 10,294 | National, regional, provincial |
| | DHS 2011 | 13,745 | 10,938 | National, regional, provincial |
| | AIS 2015 | 6,946 | 2,837 | National, regional, provincial |
| Tanzania | DHS 2004-05 | 10,329 | 8,554 | National, regional |
| | DHS 2010 | 10,139 | 7,989 | National, regional |
| | AIS/MIS 2011-12 | 10,967 | 5,833 | National, zonal, regional |
| | DHS 2015-16 | 13,266 | 10,144 | National, zonal, regional |
| **Facility readiness scores and readiness-adjusted institutional delivery coverage** | | | | |
| Health facility assessments (HFA) | | Health facilities | Health facilities | Geographic representativeness |

| | | | | |
|---|---|---|---|---|
| | | assessed | assessed offering delivery services[1] | |
| Malawi | SPA 2013-14 | 977 | 540 | Census, all levels |
| Mozambique | SARA 2018 | 1,643 | 1,391 | Census, all levels |
| Tanzania | SPA 2014-15 | 1,188 | 951 | National, regional, facility type (level and managing authority) |
| Household surveys (HH) | | Interviewed women aged 15-49 | Most recent live births[2] | Geographic representativeness |
| Malawi | DHS 2015-16 | 24,562 | 13,399 | National, regional, district |
| Mozambique | AIS 2015 | 6,946 | 2,648 | National, regional, provincial |
| Tanzania | DHS 2015-16 | 13,266 | 7,025 | National, zonal, regional |
| **Care-seeking behaviors for newborns who died** | | | | |
| Verbal and social autopsy questionnaires (VASA) | | Newborns who died | Newborns who died whose place of death was not 'en route' | Geographic representativeness |
| Malawi | CHERG Stillbirth, Neonatal, and Child VASA questionnaire, 2013 | 320 | 318 | Selected districts (Balaka and Salima) |
| Mozambique | COMSA Stillbirth, Neonatal, Child | 403 | 400 | National |

[1] HFA analytic data sets were limited to include only facilities offering delivery and newborn care services (Malawi and Tanzania) or basic obstetric care services (Mozambique).
[2] Analyses were limited to live births to interviewed women in the five years preceding the survey for which information on place of delivery was available (not missing), except for MWI MICS 2013-14 and MWI MICS 2006 which included only live births in the preceding two years and MOZ AIS 2015 which included only live births in the preceding 32 months.

| | | | | |
|---|---|---|---|---|
| | and Adult VASA questionnaire, 2018 | | | |
| Tanzania | CHERG Stillbirth, Neonatal, and Child VASA questionnaire, 2017/18 | 228 | 226 | National |

**Figure A-1. Care-seeking itinerary for newborns' fatal illness in Malawi, Mozambique, and Tanzania.**

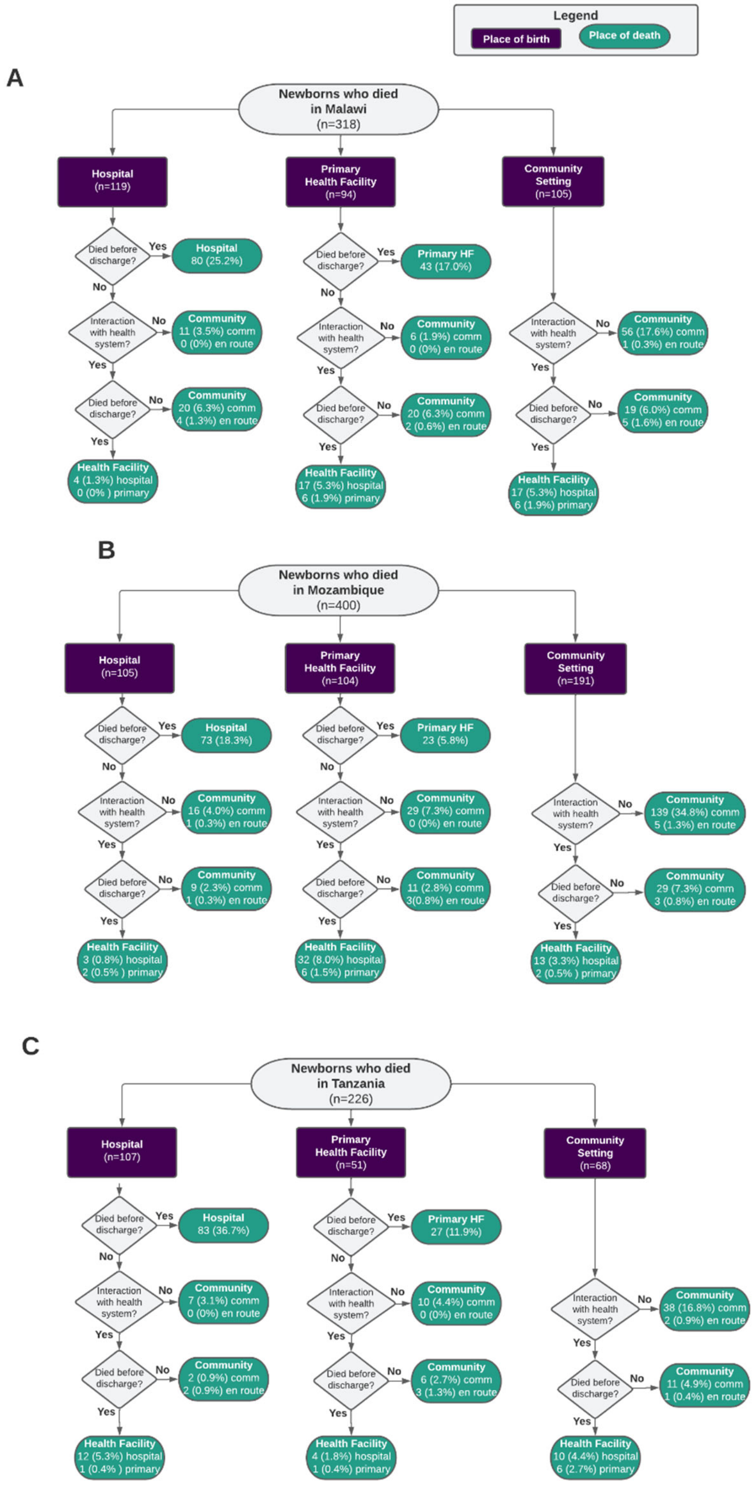


Note: Percentages are calculated by dividing the number of newborns who died in a particular place of birth and place of death category (green ovals) by the total number of newborns who died (318 in Malawi, 400 in Mozambique, and 226 in Tanzania).

**Figure A- 2 Barriers to care-seeking reported by caretakers of newborns who died in Malawi, Mozambique, and Tanzania**

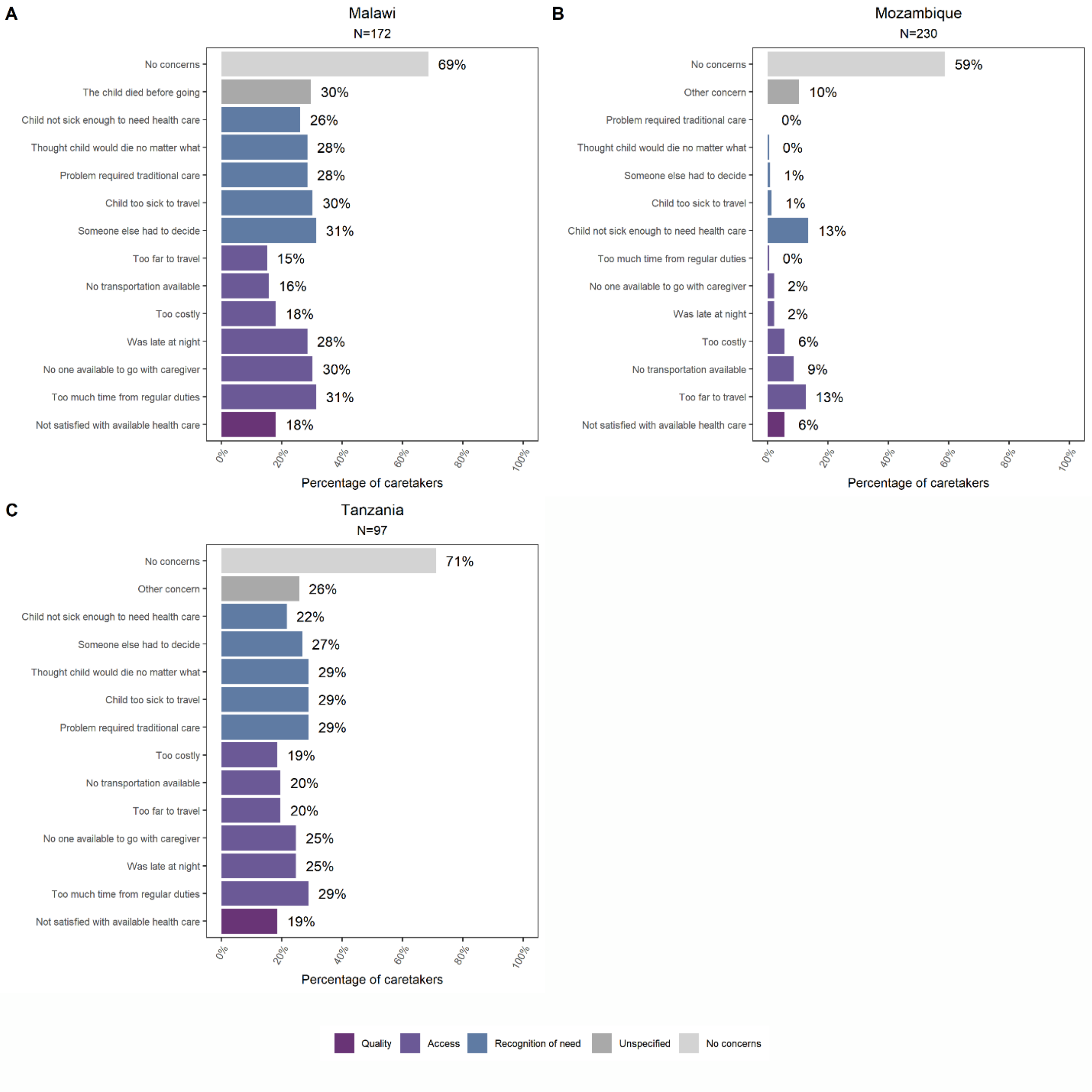

**Table A- 2. Detailed definitions and survey-specific availability for items used to measure facility readiness for each of the eight interventions required for small and sick newborn care included in the analysis.** Interventions included are essential newborn care (ENC), breastfeeding, resuscitation, prevention of mother-to-child transmission of HIV (PMTCT), kangaroo mother care (KMC), antibiotics for neonatal infection, and general readiness items (cross-cutting).

| Domain | Item | Detailed indicator definition | Data availability notes | Survey specific: Indicator not collected | Cross-country differences |
|---|---|---|---|---|---|
| **Essential newborn care (ENC)** – 9 items | | | | | |
| Equipment and supplies | Linen for drying baby | Linen for wrapping and drying the newborn observed available in the delivery service provision area. | Not collected in SARA; collected for some SPAs | MWI_2013 MOZ_2018 | |
| Equipment and supplies | Cord cutting supplies | Sterile scissors/blade to cut cord and cord clamp OR sterile delivery pack observed available in the delivery service provision area. | | | MWI and TZA were observed available while MOZ was observed available and functional |
| Equipment and supplies | Infant scale | Infant weighing scale observed available and functional in the delivery service provision area or OPD. | | | |
| Equipment and supplies | Thermometer for low-body temperature | Thermometer for low-body temperature observed available and functional in the delivery service provision area. | Not collected in SARA | MOZ_2018 | |

| | | | | | |
|---|---|---|---|---|---|
| Medicines and commodities | Vitamin K | Vitamin K observed in pharmacy or anywhere in the facility where medicines are routinely stored; at least one with valid expiration date. | Not collected in SARA; collected for some SPAs | MWI_2013 MOZ_2018 | |
| Medicines and commodities | Antibiotic eye ointment for newborns (tetracycline or other) | Antibiotic eye ointment for newborns observed in pharmacy or anywhere in the facility where medicines are routinely stored; at least one with valid expiration date. | | | |
| Medicines and commodities | Chlorhexidine solution[3] | Chlorhexidine solution observed in pharmacy or anywhere in the facility where medicines are routinely stored; at least one with valid expiration date. | Not collected in SARA | MOZ_2018 | Not collected in MOZ; CHX not part of national policy for TZA so excluded for TZA |
| Trained staff | Staff trained in clean cord cutting and appropriate cord care | Proportion of health care workers delivering newborn care services that have been trained in clean cord cutting and appropriate cord care in the last two years. | Not collected in SARA | MOZ_2018 | |
| Trained staff | Staff trained in thermal care | Proportion of health care workers delivering newborn care services that have been trained in thermal care (including immediate drying and skin-to-skin care) in the last two years. | Not collected in SARA | MOZ_2018 | |
| **Breastfeeding** – 2 items | | | | | |

[3] Only expected in countries for which chlorhexidine is part of the national policy.

| | | | | | |
|---|---|---|---|---|---|
| Trained staff | Staff trained in early and exclusive breastfeeding | Proportion of health care workers delivering newborn care services that have been trained in early and exclusive breastfeeding in the last two years. | Not collected in SARA | MOZ_2018 | |
| Routine services | Initiation of breastfeeding within the first hour | Facility routinely practices initiation of breastfeeding within the first hour. | | | |
| **Resuscitation** – 5 items | | | | | |
| Equipment and supplies | Airway suction apparatus (suction apparatus with catheter or suction bulb for mucus extraction) | Airway suction apparatus (suction apparatus with catheter or suction bulb for mucus extraction) observed available and functional in the delivery service provision area. | | | |
| Equipment and supplies | Infant resuscitation bag/mask | Infant resuscitation bag/mask observed available and functional in the delivery service provision area. | | | |
| Equipment and supplies | Stethoscope | Stethoscope observed available and functional in the delivery service provision area. | | | MOZ used item in the OPD as not collected in the delivery service area |
| Trained staff | Staff trained in neonatal resuscitation using bag and mask | Proportion of health care workers delivering newborn care services that have been trained in neonatal resuscitation using bag and mask in the last two years. | | | MOZ used alternative definition while SPA countries |

| | | | | | |
|---|---|---|---|---|---|
| | | *Alternative:* At least one health care worker providing delivery services has been trained in neonatal resuscitation using bag and mask in the last two years. | | | used preferred definition |
| Routine services | Facility past three months provided neonatal resuscitation | Providers have carried out neonatal resuscitation as part of their work in the facility in the past three months. | | | MOZ definition is past 12 months |
| **Prevention of mother-to-child transmission of HIV (PMTCT)** – 8 items | | | | | |
| Infrastructure | PMTCT room is private room with auditory and visual privacy | PMTCT room is private room with auditory and visual privacy. | | | MWI and TZA have option for PMTCT room within delivery area or as a separate service area; MOZ only asks as a separate service area; SPA countries were observed while MOZ/SARA was reported |
| Medicines and commodities | Antiretrovirals for babies | Antiretrovirals for babies (NVP or AZT syrup only) observed in pharmacy or anywhere in the facility where medicines are routinely stored; at least one with valid expiration date. | | | |

| | | | | | |
|---|---|---|---|---|---|
| Medicines and commodities | Antiretrovirals for mothers | Antiretrovirals for mothers (country-specific first line ARV prophylaxis for HIV-positive pregnant women) observed in pharmacy or anywhere in the facility where medicines are routinely stored; at least one with valid expiration date. | | | TZA: TDF/3TC/EFV or AZT/3TC/NVP MWI: TDF/3TC/EFV OR AZT/3TC/NVP MOZ: TDF/3TC/EFV or AZT/3TC/EFV or TDF/3TC/LPV) |
| Diagnostics | HIV diagnostic capacity | Facility has the ability to conduct HIV testing onsite and has the required equipment available and functioning/unexpired (RDT, ELISA (scanner/reader, washer, assay, and incubator), dynabeads, or Western blot ). | | | MWI did not ask expiration of western blot or availability of ELISA assay; MOZ did not ask Western blot test or dynabeads |
| Guidelines | PMTCT guidelines | Guidelines for PMTCT are available in the PMTCT service provision area. | | | |
| Guidelines | Infant and young child feeding counseling guidelines | Guidelines for infant and young child feeding are observed available within the PMTCT service provision area. | | | |
| Trained staff | Staff trained PMTCT or ARV prophylactic | Proportion of health care workers delivering PMTCT services that have been trained in PMTCT or ARV | | | MOZ used alternative definition while |

| | | | | | |
|---|---|---|---|---|---|
| | treatment for PMTCT | prophylactic treatment for PMTCT in the last two years.<br>Alternative: At least one health care worker providing PMTCT services has been trained in PMTCT in the last two years. | | | SPA countries used preferred definition |
| Trained staff | Staff trained newborn nutrition counseling of mother with HIV or infant and young child feeding | Proportion of health care workers delivering PMTCT services that have been trained in infant and young child feeding in the last two years.<br>Alternative: At least one health care worker providing PMTCT services has been trained in infant and young child feeding in the last two years. | | | MOZ used alternative definition while SPA countries used preferred definition |
| **Kangaroo mother care (KMC)** – 3 items | | | | | |
| Infrastructure | Separate room or space for KMC | Facility has a separate room or space for KMC. | Not collected in SARA | MOZ_2018 | |
| Trained staff | Staff trained in KMC for low-birth-weight babies | Proportion of health care workers delivering newborn care services that have been trained in KMC for low-birth-weight babies in the last two years. | Not collected in SARA | MOZ_2018 | |
| Routine services | Facility practices KMC | Facility practices KMC. | | | MOZ SARA specifies "has practiced in the last 12 months" whereas MWI and TZA SPA more broadly asks "Does the facility |

| | | | | | |
|---|---|---|---|---|---|
| | | | | | practice KMC?" |
| **Antibiotics for neonatal infection** – 3 items | | | | | |
| Equipment and supplies | Medication delivery mechanism (infusion kit + any IV fluid OR single use syringes) | Medication delivery mechanism (infusion kit + any IV fluid OR single use syringes) observed anywhere in the facility where supplies are routinely stored. | | | |
| Medicines and commodities | Antibiotic treatment for neonatal infection | Antibiotic treatment for neonatal infection (Outpatient treatment: Gentamycin (IM or IV) and Amoxicillin (syrup/oral suspension) OR Inpatient treatment: (Benzylpenicillin (IV or IM) OR Ampicillin (IV or IM)) AND Gentamycin (IM or IV)) observed in pharmacy or anywhere in the facility where medicines are routinely stored; at least one with valid expiration date. | | | |
| Trained staff | Staff trained in newborn infection management (including injectable antibiotics) | Proportion of health care workers delivering newborn care services that have been trained in newborn infection management (including injectable antibiotics) in the last two years. | Not collected in SARA | MOZ_2018 | |
| **General readiness items** – 7 items | | | | | |
| Equipment and supplies | Thermometer | Thermometer observed available and functional in the delivery service provision area or OPD. | | | MOZ used item in the OPD only as not collected |

| | | | | | |
|---|---|---|---|---|---|
| | | | | | in the delivery service area |
| Equipment and supplies | Pedal bin waste receptacle with lid and plastic liner | Pedal bin waste receptacle with lid and plastic liner observed available in the delivery service provision area. | | | MOZ used item in the OPD as not collected in the delivery service area |
| Equipment and supplies | Environmental disinfectant | Disinfectant (e.g., chlorine, hibitane, alcohol) is observed available in the delivery service provision area or OPD. | | | MOZ used item in the OPD only as not collected in the delivery service area |
| Equipment and supplies | Clean/sterile gloves | Disposable latex gloves are observed available in the delivery service provision area. | | | |
| Equipment and supplies | Sharps container | Sharps container observed available in the delivery service provision area. | | | MOZ used item in the OPD as not collected in the delivery service area |
| Equipment and supplies | Soap and water for handwashing | Handwashing soap and running water OR alcohol-based hand rub observed available in the delivery service provision area. | | | MOZ used item in the OPD as not collected in the delivery service area |
| Guidelines | Guidelines containing information on | Guidelines containing information on newborn care (e.g., ENC, IMPAC, BEmOC, CEmOC) are observed | | | TZA: BEmOC, CEmOC, newborn care |

| | | | | | |
|---|---|---|---|---|---|
| | newborn care (e.g., ENC, IMPAC, BEmOC, CEmOC) | available in the delivery service provision area. | | | guidelines<br>MWI: BEmOC, CEmOC, IMPAC<br>MOZ: ENC, essential childbirth care |

**Table A- 3. Average health facility readiness scores for items required for small and sick newborn care, by intervention and country.** Scores reflect the mean across all facility types included in each country's most recent HFA.

| SSNC Interventions | Item Number | Readiness Item | Readiness Scores | | |
|---|---|---|---|---|---|
| | | | Malawi | Mozambique | Tanzania |
| Essential Newborn Care (ENC) | 1 | Linen for drying baby | NA | NA | 0.12 |
| | 2 | Cord cutting supplies | 0.95 | 0.95 | 0.92 |
| | 3 | Infant scale | 0.95 | 0.95 | 0.80 |
| | 4 | Thermometer for low-body temperature | 0.08 | NA | 0.04 |
| | 5 | Vitamin K | NA | NA | 0.03 |
| | 6 | Antibiotic eye ointment for newborns (Tetracycline or other) | 1.00 | 0.93 | 0.51 |
| | 7 | Chlorhexidine solution | 0.36 | NA | NA |
| | 8 | Staff trained in clean cord cutting and appropriate cord care | 0.43 | NA | 0.43 |
| | 9 | Staff trained in thermal care | 0.43 | NA | 0.42 |
| Breastfeeding | 10 | Staff trained in early and exclusive breastfeeding | 0.36 | NA | 0.40 |
| | 11 | Initiation of breastfeeding within the first hour | 0.99 | 0.99 | 0.98 |
| Resuscitation | 12 | Airway suction apparatus (suction apparatus with catheter or suction bulb for mucus extraction) | 0.91 | 0.17 | 0.74 |
| | 13 | Infant resuscitation bag/mask | 0.89 | 0.76 | 0.76 |
| | 14 | Stethoscope | 0.78 | 0.80 | 0.72 |
| | 15 | Staff trained in neonatal resuscitation using bag and mask | 0.52 | 0.42 | 0.51 |
| | 16 | Facility past three months provided neonatal resuscitation | 0.88 | 0.85 | 0.52 |
| Prevention of mother-to-child transmission of HIV (PMTCT) | 17 | PMTCT room is a private room with auditory and visual privacy | 0.79 | 0.64 | 0.73 |
| | 18 | HIV diagnostic capacity | 0.97 | 0.99 | 0.91 |
| | 19 | Antiretrovirals for babies | 0.87 | 0.79 | 0.59 |
| | 20 | Antiretrovirals for mothers | 0.97 | 0.83 | 0.80 |
| | 21 | PMTCT guidelines | 0.57 | 0.72 | 0.70 |
| | 22 | IYCF counselling guidelines | 0.33 | 0.46 | 0.32 |
| | 23 | Staff trained in PMTCT or ARV prophylactic treatment for PMTCT | 0.38 | 0.48 | 0.36 |
| | 24 | Staff trained in newborn nutrition counseling of mother with HIV or IYCF | 0.31 | 0.37 | 0.29 |
| Kangaroo mother care (KMC) | 25 | Separate room or space for KMC | 0.16 | NA | 0.04 |
| | 26 | Staff trained in KMC for LBW babies | 0.29 | NA | 0.32 |
| | 27 | Facility practices KMC | 0.56 | 0.83 | 0.21 |
| | 28 | Antibiotic treatment for neonatal infection | 0.89 | 0.68 | 0.28 |

| | | | | | |
|---|---|---|---|---|---|
| **Antibiotics for neonatal infection** | **29** | Medication delivery mechanism (infusion kit + any IV fluid OR single use syringes) | 0.99 | 1.00 | 0.92 |
| | **30** | Staff trained in newborn infection management (including injectable antibiotics) | 0.28 | NA | 0.30 |
| **General readiness items** | **31** | Thermometer | 0.84 | 0.85 | 0.64 |
| | **32** | Pedal bin waste receptacle with lid and plastic liner | 0.47 | 0.66 | 0.55 |
| | **33** | Environmental disinfectant | 0.84 | 0.75 | 0.78 |
| | **34** | Clean/sterile gloves | 0.97 | 0.87 | 0.86 |
| | **35** | Sharps container | 0.97 | 0.96 | 0.93 |
| | **36** | Soap and water for handwashing | 0.75 | 0.82 | 0.67 |
| | **37** | Guidelines containing information on newborn care | 0.66 | 0.58 | 0.66 |

**Table A- 4. Average overall SSNC readiness score and readiness-adjusted institutional delivery coverage, by facility type in Malawi, Mozambique, and Tanzania.**

| Facility Type | Overall SSNC Mean Facility Readiness Score | Readiness-adjusted institutional delivery coverage (95% CI) |
|---|---|---|
| **Malawi** | | |
| Home | 0 | 0.086 (0.076-0.081) |
| Public - hospital | 0.73 | 0.274 (0.256-0.265) |
| Public - health centre | 0.64 | 0.503 (0.482-0.493) |
| Public - health post | 0.62 | 0.011 (0.008-0.009) |
| CHAM - hospital | 0.74 | 0.057 (0.045-0.051) |
| CHAM - health centre/post | 0.62 | 0.048 (0.039-0.044) |
| Private - any type | 0.60 | 0.022 (0.016-0.019) |
| **Mozambique** | | |
| Home | 0 | 0.302 (0.265-0.284) |
| Public - hospital | 0.90 | 0.225 (0.198-0.212) |
| Public - primary | 0.80 | 0.469 (0.43-0.45) |
| Private - any type | 0.75 | 0.004 (0.001-0.003) |
| **Tanzania** | | |
| Home | 0 | 0.374 (0.347-0.361) |
| Public - natl hospital | 0.67 | 0.016 (0.013-0.015) |
| Public - regional hospital | 0.73 | 0.099 (0.085-0.092) |
| Public - district hospital | 0.70 | 0.126 (0.111-0.118) |
| Public - health centre | 0.61 | 0.121 (0.107-0.114) |
| Public - dispensary/clinic | 0.51 | 0.144 (0.129-0.137) |
| Mission - hospital | 0.72 | 0.071 (0.059-0.065) |
| Mission - health centre | 0.66 | 0.017 (0.012-0.015) |
| Mission - dispensary/clinic | 0.54 | 0.007 (0.005-0.006) |
| Private - hospital | 0.63 | 0.012 (0.009-0.011) |
| Private - health centre | 0.57 | 0.005 (0.003-0.004) |
| Private - dispensary | 0.43 | 0.006 (0.003-0.005) |
| Private - clinic | 0.56 | 0.001 (0-0) |
